# Combined Effects of Severe Immunocompromise and Prolonged Virus Shedding on Within-Host SARS-CoV-2 Evolution in COVID-19

**DOI:** 10.64898/2026.04.14.26350918

**Authors:** Yuichiro Hirata, Kenichiro Takahashi, Noriko Iwamoto, Yong Dam Jeong, Sho Miyamoto, Junna Kawasaki, Montie Tadaharu Harrison, Sohtaro Mine, Shun Iida, Shinji Saito, Akira Ainai, Takayuki Kanno, Harutaka Katano, Naobumi Sasaki, Kazuhiro Horiba, Takeru Tsunoi, Masahiro Ishikane, Kohei Kamegai, Naoya Itoh, Nana Akazawa-Kai, Nobumasa Okumura, Mizuki Haraguchi, Takashi Sakoh, Masayo Morishima, Hideki Araoka, Naoyuki Uchida, Ryota Hase, Yoshiaki Marumo, Takuya Adachi, Kosei Matsue, Tomoya Saito, Norio Ohmagari, Shingo Iwami, Tadaki Suzuki

## Abstract

**Background:** Prolonged SARS-CoV-2 infection in immunocompromised individuals may accelerate virus evolution within the host, raising concerns about the virus evading immunity, developing resistance, and forming novel variants of concern. However, the determinants and public health implications of within-host viral evolution in this population remain incompletely understood.

**Methods:** We performed longitudinal analyses of SARS-CoV-2 genomes from 91 patients with COVID-19 who were classified as being severely or moderately immunocompromised. Using serial measurements of viral RNA loads and infectious titers, we modeled the shedding dynamics of the virus and stratified the infected cases by upper respiratory virus shedding duration to assess associations with within-host evolutionary dynamics.

**Results:** Shedding modeling identified two profiles of shedding duration: intermediate and long. The long shedding profile (shedding lasting >21 days) was found in 14.8% of moderately immunocompromised cases and 72.1% of severely immunocompromised cases. Frequent single-nucleotide variants accumulated specifically in severely immunocompromised individuals with the long shedding phenotype, correlating positively with shedding duration. By contrast, mutations remained limited in moderately immunocompromised individuals with the long shedding phenotype and in severely immunocompromised individuals with the intermediate shedding phenotype. We identified mutations in the spike receptor-binding domain associated with monoclonal antibody resistance; however, we found no fitness-enhancing mutations for inter-host transmission, and antiviral drug resistance mutations were rare. Instead, mutations were introduced frequently and randomly across the entire viral genome.

**Conclusions:** Prolonged upper respiratory virus shedding exceeding 21 days combined with severe immunocompromise is a risk factor of the accumulation of within-host SARS-CoV-2 mutations. Although no variants of concern emerged, the introduction of genome-wide random mutations suggests that the risk for novel variant generation cannot be excluded. These findings highlight the need for intensive antiviral strategies to limit shedding duration to less than 21 days in severely immunocompromised patients, and for immunological investigations to elucidate the host factors underlying residual shedding control in those who achieve clearance within this threshold.

## Introduction

Since the onset of the SARS-CoV-2 pandemic, the management of COVID-19 in immunocompromised individuals, specifically people whose immunocompromised state results from underlying disease or immunosuppressive treatment, has remained a persistent clinical challenge. Immunocompromised individuals are at greater risk of developing severe COVID-19, of requiring intensive care, and of experiencing in-hospital mortality than are immunocompetent individuals^1^. Notably, this population has been observed to experience prolonged shedding of SARS-CoV-2 from the upper respiratory tract, which often leads to recurrent pneumonia and progression to respiratory failure^2^.

Prolonged virus shedding in immunocompromised hosts frequently involves extensive evolution of the virus within the host. Multiple reports have documented increased mutation rates during prolonged virus shedding^3,4^, and therapeutic interventions such as monoclonal antibodies and antiviral agents have been shown to promote the emergence of mutations that allow the virus to evade the immune system or develop drug resistance^5-7^. Moreover, such mutations may have a role in the emergence of novel variants of concern (VOCs)^8,9^. The sudden appearance of highly mutated VOCs— such as Alpha, Delta, and Omicron—each of which harbors numerous mutations relative to previously circulating lineages, led to the hypothesis that immunocompromised individuals may serve as reservoirs for accelerated viral evolution^10-12^. However, the key determinants of when and to what extent such evolution occurs remain incompletely understood.

Despite numerous case reports and viral genome sequencing studies describing within-host mutations in immunocompromised patients, as well as several cohort-based investigations^13,14^, the fundamental determinants of within-host viral evolutionary dynamics remain poorly defined. In particular, the current classifications based on clinical variables in an immunocompromised patient do not fully capture the heterogeneity in viral persistence and evolutionary potential across individuals. This is particularly relevant in the Omicron era, because the strains circulating at the population level have already acquired key mutations associated with immune evasion and drug resistance^15^. This background makes it increasingly difficult to distinguish clinically or epidemiologically meaningful within-host evolution from background genetic variation. As a result, genomic analyses of within-host mutations have largely remained descriptive, with limited ability to stratify patients according to their risk of prolonged virus shedding or to identify the contexts in which viral evolution is most likely to occur.

To address this gap, we assembled a longitudinal cohort of immunocompromised patients with COVID-19 and performed repeated virological and genomic analyses of upper respiratory specimens throughout the course of infection. A critical methodological challenge in such analyses is being able to accurately estimate the duration of virus shedding in individual cases. In immunocompromised patients with prolonged virus shedding, the observation period is necessarily extended and inter-individual variability in shedding dynamics is substantial. Because frequent specimen collection at short intervals imposes a considerable burden on patients, sampling in routine clinical practice is inevitably sparse and irregularly timed, introducing significant bias when shedding duration is inferred directly from observed data alone. To overcome this limitation, we applied mathematical modeling to serial measurements of viral RNA load and culture-based infectious virus titers. This modeling allowed us to robustly estimate the duration of virus shedding from the upper respiratory tract in each individual case. We then used the individually modeled shedding duration to examine the determinants of within-host virus evolution. We found that a prolonged period of virus shedding from the upper respiratory tract exceeding 21 days, rather than immunocompromised status alone, is the risk factor associated with increased viral genome mutation burden. These results provide a mechanistic framework linking prolonged virus shedding to SARS-CoV-2 within-host evolution and clarify the clinical contexts in which within-host mutations are most likely to arise^16^.

## Methods

### Study design (Supplementary Figure S1)

Patients with COVID-19 and underlying immunocompromised conditions were enrolled from July 2021 through January 2024 if they met the following criteria: (1) laboratory-confirmed SARS-CoV-2 infection by polymerase chain reaction (PCR), loop-mediated isothermal amplification (LAMP), or rapid antigen testing and (2) moderate to severe immunocompromised status. Immunocompromised conditions included active treatment for solid tumors or hematologic malignancies, human immunodeficiency virus infection (CD4 < 200/mm³), primary immunodeficiency, solid organ or hematopoietic stem cell transplants, active autoimmune diseases, or a history of immunosuppressant use within the past 3 months.

According to the study protocol, nasopharyngeal swabs were collected from registered patients on the day of admission; on days 5 (± 1), 10 (± 2), and 20 (± 2) of admission; at discharge; and approximately 1 month after discharge. Blood samples were collected at the following time points: at admission, at discharge, and approximately 1 month after discharge. Additional specimens obtained according to clinical indications were also included. Samples were processed for nucleic acid extraction and subjected to reverse transcription quantitative polymerase chain reaction (RT-qPCR) and viral genome sequencing. Viral isolation assays were conducted separately using the original clinical specimens.

When genomic analysis demonstrated that sequential hospitalization episodes were caused by viruses belonging to the same lineage, the episodes were classified as a single case of recurrence. In contrast, when the interval between hospitalization episodes exceeded 90 days and the viruses belonged to different lineages, the episodes were defined as separate cases of reinfection^17^.

Clinical information was collected in a standardized format, including demographic characteristics (sex, age, race, and body mass index), comorbidities, clinical course, vaccination status, and treatment.

For comparative analyses, previously reported immunocompetent patients with COVID-19 enrolled from December 2021 through January 2022 were included as a reference cohort. Corresponding virological data were incorporated from a previously published study^18^.

### Classification of immunocompromised conditions

Immunocompromised patients were classified as being severely or moderately immunocompromised according to the indication for tixagevimab/cilgavimab from the Japanese national COVID-19 guidelines^19^. The detailed criteria and patient-level classification are summarized in Supplementary Table S1. Patients not meeting the predefined criteria for severe immunocompromise were categorized as being moderately immunocompromised.

### Nucleic acid extraction and detection of SARS-CoV-2 by RT-qPCR

Nucleic acids were extracted from 200 µL of nasopharyngeal swab samples using either the Maxwell RSC miRNA Plasma and Serum Kit (Promega, Madison, WI) or the MagMAX Viral/Pathogen Nucleic Acid Isolation Kit (Thermo Scientific, Waltham, MA). Extracted nucleic acids were subjected to RT-qPCR using the One Step PrimeScript™ III RT-qPCR Mix (Takara Bio, Shiga, Japan), targeting the SARS-CoV-2 nucleoprotein (N) gene20. RT-qPCR was performed on a LightCycler 96 (Roche, Basel, Switzerland) under the following reaction conditions: 52°C for 5 min, 95°C for 10 s, followed by 45 cycles of 95°C for 5 s and 60°C for 30 s. Quantification cycle (Cq) values were converted to viral RNA copy numbers per reaction based on a simple regression line^21^.

### SARS-CoV-2 virus isolation assay

Virus isolation was performed as previously described^20^. Clinical specimens were diluted in Dulbecco’s modified Eagle medium supplemented with antibiotic-antimycotic 2% fetal bovine serum (Thermo Fisher Scientific, MA, USA) and inoculated onto VeroE6/TMPRSS2 cells (JCRB1819, JCRB Cell Bank, Osaka, Japan) seeded in 96-well flat-bottom plates. Culture supernatant was replaced 1 day post infection (dpi), and cells were incubated at 37 ℃ with 5% CO_2_. Cytopathic effects (CPE) were observed at 1, 4, and 5 dpi. At 5 dpi, culture supernatants were collected and subjected to RT-qPCR using the SARS-CoV-2 Direct Detection RT-qPCR Kit (Takara Bio, Shiga, Japan) to confirm virus replication. A sample was considered virus-isolation-positive when both CPE and RT-qPCR positivity were observed. Infectious virus titers were quantified as the median tissue culture infectious dose (TCID₅₀) for isolation-positive samples.

### Characterizing SARS-CoV-2 shedding dynamics from serial RNA and infectious titers

We used a mathematical modeling framework to analyze longitudinal measurements of viral RNA loads and infectious virus titers to characterize within-host virus dynamics. The modeling cohort was defined by the availability of repeated RNA/titer measurements and partially overlapped with the cohort used for genome analyses. The model links RNA kinetics to infectious virus dynamics and allows us to estimate patient-specific shedding characteristics, including the time to RNA negativity and the time to culture negativity. To further capture heterogeneity among immunocompromised patients, we used these model-estimated shedding features as inputs for hierarchical clustering. This method identified two virologically distinct groups, one with an intermediate duration of shedding and one with a long duration of shedding. Detailed modeling and clustering procedures are described in the Supplementary Methods.

### SARS-CoV-2 whole viral genome sequencing

Samples with sufficient SARS-CoV-2 viral load (generally ≥10⁵ RNA copies/mL) were subjected to whole-genome sequencing. Five microliters of the extracted nucleic acid was used for library propagation according to the nCoV-2019 sequencing protocol for Illumina v5^21^. Multiplex RT-qPCR amplification was performed using specifically designed primers^22^, and the amplicons were processed with the QIAseq FX DNA Library Kit (Qiagen, Hilden, Germany). Libraries were sequenced on MiSeq or NextSeq 1000 platforms (Illumina, San Diego, CA, USA).

Sequencing reads were mapped to the SARS-CoV-2 reference genome MN908947.3^23^ using bwa v. 0.7.13-r1126^24^, and variant calling was performed using GATK v. 3.8.0^25^ and VarScan v. 2.4.3^26^. Upon integrating the variant call data and *de novo* assemblies produced by SKESA v. 2.3.0^27^ or A5-miseq v. 20140604^28^, consensus sequences were generated^29^. Manual curation and Sanger sequencing were additionally used to resolve ambiguous regions where necessary.

The sequencing data have been deposited in the DDBJ Sequence Read Archive under BioProject accession PRJDB20632, with Run accessions DRR667741–DRR668026 (Supplementary Dataset).

### Nucleotide and amino acid mutation analysis and phylogenetic analysis

Consensus viral genome sequences were analyzed to identify nucleotide and amino acid substitutions and to assign Pango lineages using Nextclade v3.13.0^30,31^. Multiple sequence alignment was performed using MAFFT v7.525^32^. Phylogenetic trees were constructed using the maximum-likelihood method under the Kimura 2-parameter (K80) model^33^. All phylogenetic analyses were performed using MEGA12^34^, and trees were visualized using FigTree v1.4.4.

### Amino acid mutations associated with resistance to therapeutic monoclonal antibodies

Amino acid substitutions located within the receptor-binding domain (RBD) and previously reported to confer resistance to therapeutic monoclonal antibodies (mAbs) were identified and analyzed. Therapeutic mAbs were categorized according to their binding sites or functional mechanisms as previously described^35^. The association between the emergence of resistance-associated substitutions and the administration of corresponding mAbs was assessed within each therapeutic category. Resistance-associated sites were defined based on published evidence for tixagevimab, casirivimab, cilgavimab, imdevimab, and sotrovimab (Supplementary Table S2). Structural visualization of substitution sites was performed using 6M0J^36^ from the Protein Data Bank and UCSF ChimeraX version: 1.8^37^.

### Analysis of SARS-CoV-2 community transmissibility using a protein language model

Amino acid sequences of the SARS-CoV-2 spike protein were extracted from viral genome sequences using Nextclade CLI v.3.8.2^31^. Transmissibility was evaluated using CoVFit, a computational framework based on protein language models that estimates relative fitness from spike protein sequences^16^.

For contextual comparison, contemporaneous SARS-CoV-2 genome sequences from Japan were retrieved from the Global Initiative on Sharing All Influenza Data (GISAID) and filtered according to predefined quality criteria (Supplementary Methods). Fitness scores were calculated for sequences derived from immunocompromised patients (classified as having severe or moderate immunocompromise) and for community sequences representing the general population. The resulting fitness scores were used as a proxy for relative community transmissibility.

### In-depth phylogenetic and haplotype network analyses of prolonged infection

Phylogenetic and haplotype network analyses were performed for CID-050, the case with the longest genome observation period in this cohort, to further characterize within-host viral evolution.

Maximum-likelihood phylogenetic trees were constructed using IQ-TREE multicore version 2.4.0^38^ with 1,000 bootstrap replicates. The best-fit substitution model (TN+F+I) was selected according to the Bayesian Information Criterion. Bayesian phylogenetic analysis was performed using BEAST2 v2.7.7^39^ based on consensus viral genome sequences and their corresponding collection dates under a strict molecular clock model. Markov chain Monte Carlo (MCMC) was run for 10,000,000 steps, and effective sample sizes (ESS) >200 were confirmed for key parameters, including the substitution rate (clockRate). The evolutionary rate was estimated from the posterior distribution. A maximum clade credibility (MCC) tree was generated after discarding burn-in. Trees were visualized using R (version 4.5.0).

Haplotype network analysis was performed using PopART (version 1.7) ^40^ with the median-joining network method.

### Electrochemiluminescence immunoassay

Serum specimens were heat-treated at 56°C for 30 minutes prior to antibody analysis. Antibody titers for the ancestral spike (S) receptor-binding domain were measured using the Elecsys Anti-SARS-CoV-2 S kits (Roche, Basel, Switzerland), according to the manufacturer’s instructions.

### Statistical analysis and reproducibility

Comparisons between two groups were performed using the Mann-Whitney U test. Comparisons among multiple groups were conducted using the Kruskal-Wallis test, followed by Dunn’s multiple-comparisons test with adjustment for multiple testing. Correlations between variables were assessed using Spearman’s rank correlation coefficient. Differences in slopes between groups were evaluated using linear regression models with group-by-variable interaction terms. Fisher’s exact test was used to assess associations between categorical variables. Nonlinear regression using a Gaussian model was applied to model data distribution. All statistical tests were two-sided, and a P value < 0.05 was considered statistically significant. Statistical analyses were performed using GraphPad Prism 9.5.1 (GraphPad Software, San Diego, CA, USA) or R (version 4.5.1).

### Ethical approval

This study was approved by the Medical Research Ethics Committee of the National Institute of Infectious Diseases (NIID, approval no. 1821). Written informed consent was obtained from all participants or their legal guardians.

The use of viral genome sequence data from a Japanese epidemiological study conducted during the Omicron wave was separately approved by the NIID Ethics Committee (approval no. 1940).

## Results

### Patient demographics and clinical characteristics

During the study period, samples were obtained from 91 immunocompromised patients with COVID-19 (Case ID: CID-XXX, Supplementary Dataset). Two patients experienced reinfection and were therefore analyzed as separate infection cases (CID-024-1, -2, and CID-044-1, -2). After we applied the predefined exclusion criteria, 88 cases were included in the final analysis (Supplementary Figure S1). Among these, 53 cases were classified in the severely immunocompromised group, and 35 cases were classified in the moderately immunocompromised group. Two of the reinfected cases (CID-024-1 and CID-024-2) were classified in the severely immunocompromised group. In contrast, CID-044-1 was categorized in the moderately immunocompromised group, whereas CID-044-2 was categorized in the severely immunocompromised group owing to a change in the patient’s condition.

The distribution of underlying comorbidities contributing to the patients’ immunocompromised status differed between the moderately and severely immunocompromised groups (Table 1). In the moderately immunocompromised group, solid cancer and recent chemotherapy were the primary contributing factors. In contrast, the severely immunocompromised group predominantly consisted of patients with hematologic malignancies. Notably, hematologic malignancies and associated B-cell-depleting therapy were key criteria for eligibility for tixagevimab/cilgavimab monoclonal antibody therapy (Supplementary Table S1).

**Table 1.** Demographic Data and Clinical Information According to Immunocompromised Status.

|  | Moderate (n = 35) | Severe (n = 53) |
| --- | --- | --- |
| Sex |  |  |
| Men | 19 (54%) [38–70%] | 35 (66%) [53–77%] |
| Age, years | 68 [64–72] | 62 [58–67] |
| Race |  |  |
| Japan | 35 (100%) [90–100%] | 49 (94%) [84–98%] |
| Asia (non-Japan) | 0 [0–10%] | 2 (4%) [1–13%] |
| Unknown | 0 [0–10%] | 1 (2%) [0–10%] |
| BMI | 22.9 [21.5–24.3] | 21.2 [20.3–22.2] * |
| Comorbidities |  |  |
| Solid cancer | 31 (89%) [74–95%] | 4 (8%) [3–18%] |
| Hematologic malignancy | 2 (6%) [1–19%] | 48 (92%) [80–96%] |
| Autoimmune disease | 1 (3%) [0–15%] | 3 (6%) [2–15%] |
| HIV infection | 1 (3%) [0–15%] | 2 (4%) [1–13%] |
| AIDS | 0 [0–10%] | 1 (2%) [0–10%] |
| Use of immunosuppressants within the past 3 months | 6 (20%) [10–37%] # | 12 (24%) [14–37%] \$ |
| Chemotherapy within the past 3 months (excluding hormonal therapy) | 26 (87%) [70–95%] # | 14 (28%) [17–42%] \$ |
| Clinical course |  |  |
| Pneumonia on admission | 7 (20%) [10–36%] | 40 (75%) [62–85%] |
| Oxygen therapy | 11 (31%) [19–48%] | 29 (55%) [41–67%] |
| Intensive care | 1 (3%) [0–15%] | 10 (19%) [11–31%] |
| Mechanical respiratory support | 0 [0–10%] | 6 (11%) [5–23%] |
| Death | 2 (6%) [1–19%] | 10 (19%) [11–31%] |
| Number of COVID-19 vaccinations |  |  |
| 0 | 6 (17%) [8–33%] | 15 (30%) [20–44%] |
| 1 | 0 [0–10%] | 2 (4%) [1–13%] |
| 2 | 7 (20%) [10–36%] | 13 (25%) [15–38%] |
| 3 or more | 22 (63%) [46–77%] | 22 (42%) [29–55%] |
| Treatment |  |  |
| Remdesivir | 30 (86%) [71–94%] | 45 (85%) [73–92%] |
| Molnupiravir | 2 (6%) [1–19%] | 3 (6%) [2–15%] |
| Nirmatrelvir/Ritonavir | 2 (6%) [1–19%] | 23 (43%) [31–57%] |
| Ensitrelvir | 0 [0–10%] | 3 (6%) [2–15%] |
| Tixagevimab/Cilgavimab | 0 [0–10%] | 22 (42%) [29–55%] |
| Casirivimab/Imdevimab | 0 [0–10%] | 6 (11%) [5–23%] |
| Sotrovimab | 0 [0–10%] | 4 (8%) [3–18%] |
| Tocilizumab | 1 (3%) [0–15%] | 9 (17%) [9–29%] |
| Baricitinib | 1 (3%) [0–15%] | 6 (11%) [5–23%] |
| Steroids | 6 (17%) [8–33%] | 32 (60%) [47–72%] |
| SARS-CoV-2 lineages |  |  |
| Delta | 0 [0–10%] | 2 (4%) [1–13%] |
| BA.1 sublineage | 0 [0–10%] | 4 (8%) [3–18%] |
| BA.2 sublineage | 4 (11%) [5–26%] | 8 (15%) [8–27%] |
| BA.5 sublineage | 23 (66%) [49–79%] | 22 (42%) [29–55%] |
| XBB sublineage | 6 (17%) [8–33%] | 14 (26%) [16–40%] |
| Others | 1 (3%) [0–15%] | 1 (2%) [0–10%] |
| Undetermined | 1 (3%) [0–15%] | 2 (4%) [1–13%] |

Data are presented as mean [95% CI, lower limit–upper limit] or n (%) [95% CI, lower limit–upper limit]; BMI, body mass index; mAb, monoclonal antibody.
\*; One case had missing data. <sup>#</sup>; Five cases had missing data. <sup>\$</sup>; Three cases had missing data.

The severely immunocompromised group tended to exhibit more severe symptoms than the moderately immunocompromised group. In terms of pharmacologic treatment, antiviral drugs, mainly remdesivir, were used in both groups, whereas monoclonal antibodies were used in the severely immunocompromised group only. Immunosuppressants, such as steroids, were also more frequently administered in the severely immunocompromised group.

### Model-based identification of prolonged upper respiratory infectious virus shedding phenotypes

SARS-CoV-2 viral RNA shedding and infectious virus shedding were both significantly prolonged in the severely immunocompromised group (log-rank test, P < 0.0001; Figure 1a), with median times to last positive RNA detection of 10 days [95% CI: 10 – 12] in the moderately immunocompromised group and 60 days [95% CI: 29 – 81] in the severely immunocompromised group, and median times to last positive viral isolation of 5 days [95% CI: 3 – 5] and 33 days [95% CI: 18 – 57], respectively (Supplementary Table S3).

**Figure 1.**
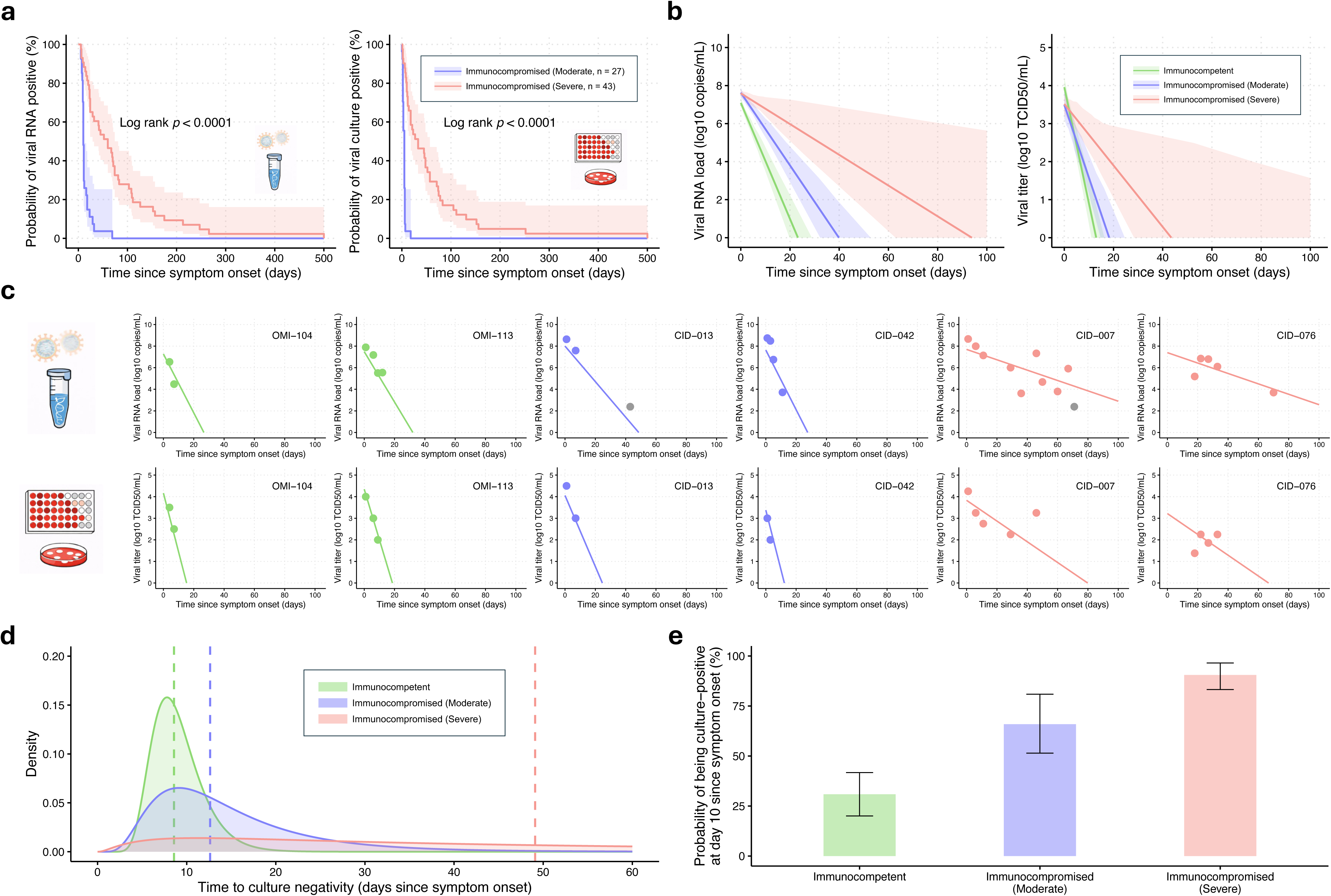
Prolonged viral RNA and culture positivity and model-based characterization of infectious shedding dynamics in immunocompromised patients. (a) Kaplan–Meier survival curves showing the probability of remaining viral RNA positive (left) and viral culture positive (right) since symptom onset in immunocompromised cases classified as Moderate (blue, n = 27) or Severe (red, n = 43). Shaded regions indicate 95% confidence intervals. P values were calculated using the log-rank test. (b) Estimated group-level trajectories of viral RNA load (left) and infectious viral titer (right) since symptom onset for the immunocompetent (green), Moderate (blue), and Severe (red) groups, based on the mathematical model fitted to longitudinal measurements. Solid lines indicate model-estimated mean trajectories, and shaded regions indicate the interquartile range (IQR; 25th–75th percentiles). (c) Representative individual-level fitted trajectories for six cases (two per group), showing viral RNA load (top) and infectious viral titer (bottom) over time since symptom onset. Solid lines indicate fitted model trajectories, and circles indicate observed measurements. (d) Fitted lognormal probability density functions of the estimated time to culture negativity (days since symptom onset) for the immunocompetent (green), Moderate (blue), and Severe (red) groups. Vertical dashed lines indicate the median of each fitted distribution. (e) Model-estimated probability of remaining culture-positive at day 10 since symptom onset for each group, calculated from the fitted lognormal distributions as 1 – F(10), where F is the cumulative distribution function (CDF). Error bars indicate 95% confidence intervals obtained via bootstrap resampling.

Using serial RNA viral load and infectious virus titers, we estimated group-level virological trajectories over time (Figure 1b and 1c). The severely immunocompromised group showed slower viral clearance than the immunocompetent and moderately immunocompromised groups (Supplementary Table S4), resulting in a longer duration of detectable viral RNA. We then inferred the duration of culture-based infectious viral shedding, summarized as time to culture negativity, and fitted a lognormal distribution to the estimated durations (Figure 1d and Supplementary Table S5). The median time to culture negativity was 8.6 days [95% CI: 7.8 – 9.4] in the immunocompetent group and 12.6 days [95% CI: 10.2 – 15.6] in the moderately immunocompromised group. By contrast, the value was 49.1 days [95% CI: 34.3 – 70.9] in the severely immunocompromised group, which exhibited a markedly right-skewed distribution with a long tail. We also estimated the probability of remaining culture-positive at day 10 since symptom onset, a time point commonly used in isolation practices during the COVID-19 pandemic. Previous studies have reported that the ability to culture SARS-CoV-2 substantially declines beyond day 10 after symptom onset^41,42^. In our analysis, however, the estimated probability remained higher in the immunocompromised groups (moderate: 65.9% [95% CI: 51.2 – 80.8]; severe: 90.5% [95% CI: 83.3 – 96.5]) than in the immunocompetent group (30.9% [95% CI: 20.0 – 41.8]) (Figure 1e).

To further characterize the heterogeneity in infectious virus shedding among immunocompromised patients, we performed a model-based reclassification based on virological rather than clinical features. Hierarchical clustering based on the estimated time to RNA negativity and time to culture negativity identified two distinct shedding phenotypes: “intermediate” (i.e., an intermediate duration of virus shedding) and “long” (i.e., a longer duration of virus shedding) (Figure 2a, Supplementary Table S6 and S7). The group with the long shedding phenotype showed significantly delayed clearance of both viral RNA and infectious virus compared with the intermediate group, whereas both groups had higher viral RNA loads at symptom onset than the group with a normal shedding duration (Figure 2b). These differences were also reflected in the Kaplan–Meier curves, in which the group with the long phenotype remained RNA-positive and culture-positive for substantially longer than did the group with the intermediate phenotype (log-rank test, P < 0.0001 for both; Figure 2c).

**Figure 2.**
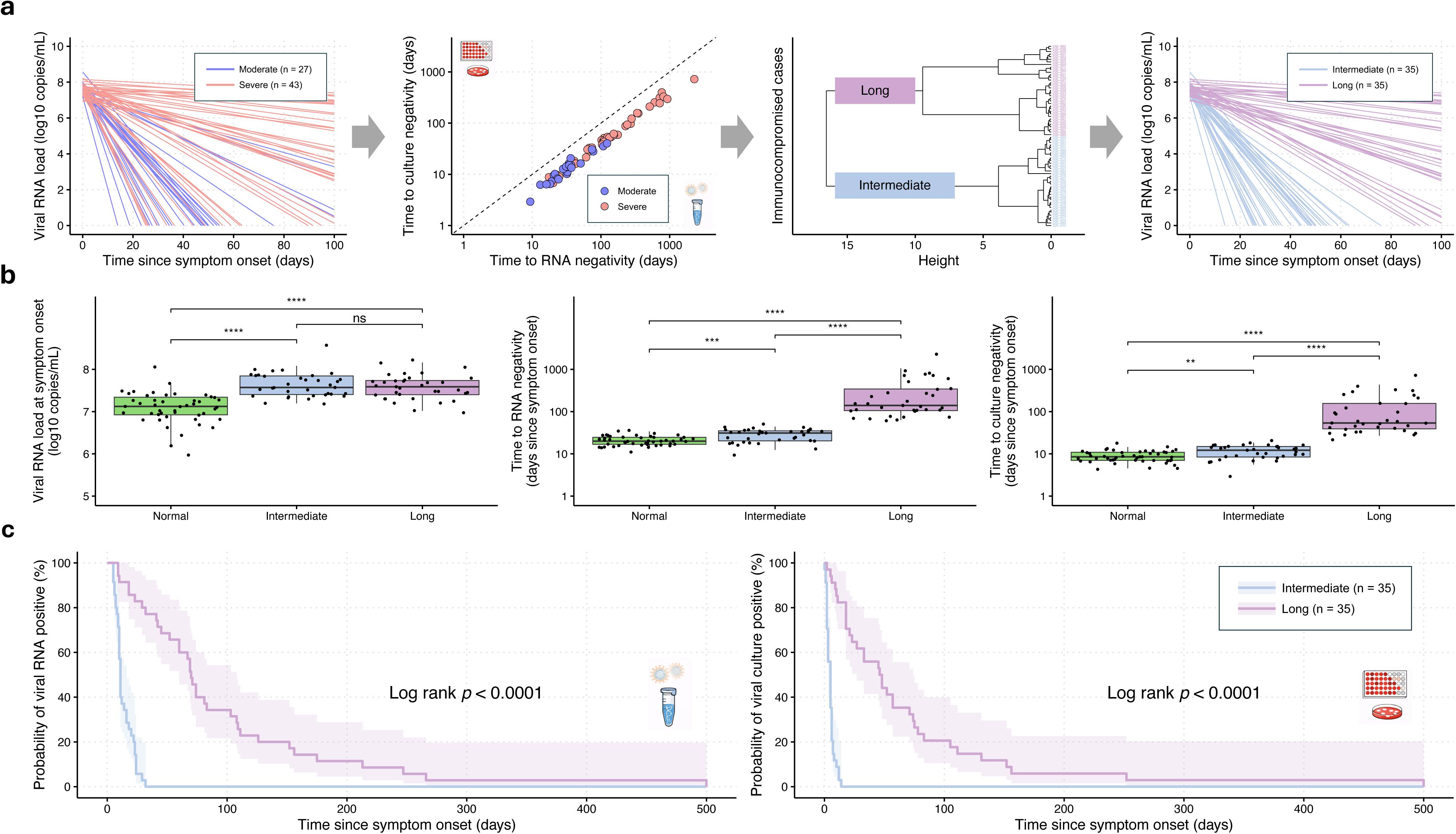
Reclassification of immunocompromised cases by shedding duration identifies distinct viral shedding patterns. (a) Schematic overview of the reclassification procedure for immunocompromised cases. Left, individual viral RNA load trajectories in immunocompromised cases clinically classified as Moderate (blue, n = 27) or Severe (red, n = 43). Middle left, two-dimensional feature space defined by the estimated time to RNA negativity and the estimated time to culture negativity for each immunocompromised case. Middle right, hierarchical clustering based on these two estimated shedding-duration features, identifying two virologically distinct groups: Intermediate shedding duration (light blue, n = 35) and Long shedding duration (purple, n = 35). Right, individual viral RNA load trajectories after reclassification into the two shedding groups. (b) Comparison of viral RNA load at symptom onset (left), time to RNA negativity (middle), and time to culture negativity (right) among the Normal, Intermediate, and Long shedding groups. Box-and-whisker plots show the medians (50th percentile; bold lines), interquartile ranges (25th and 75th percentiles; boxes), and 2.5th to 97.5th percentile ranges (whiskers). Statistical comparisons were performed using the two-sided Wilcoxon rank-sum test. ns, not significant; *** P < 0.001; **** P < 0.0001. (c) Kaplan–Meier curves showing the probability of remaining viral RNA–positive (left) and viral culture–positive (right) since symptom onset in the Intermediate (light blue, n = 35) and Long (purple, n = 35) shedding groups. Shaded regions indicate 95% confidence intervals. P values were calculated using the log-rank test.

Severely immunocompromised status was significantly associated with the long shedding phenotype (Fisher’s exact test, P < 0.0001; Table 2). Among severely immunocompromised individuals, 72.1% were classified as having the long phenotype, compared with 14.8% of moderately immunocompromised individuals. However, 27.9% of severely immunocompromised cases were classified as having the intermediate phenotype, and 85.2% of moderately immunocompromised cases were classified similarly, demonstrating that immunocompromised status alone does not fully determine shedding phenotype and that substantial heterogeneity in virus shedding dynamics exists within each category of immunocompromise.

**Table 2.** Clinical Immunocompromised Status and Model-Based Shedding Duration Classification.

|  |  | Immunocompromised status |  |
| --- | --- | --- | --- |
|  |  | Moderate (n=27) | Severe (n=43) |
| Shedding duration | Intermediate | 23 (85.2%) [67.5-94.1%] | 12 (27.9%) [16.7-42.7%] |
|  | Long | 4 (14.8%) [5.9-32.5%] | 31 (72.1%) [57.3-83.3%] |

Data are presented as n (%) [95% CI, lower limit–upper limit]. Fisher's exact test; P < 0.0001.

### Mutation burden is associated with prolonged virus shedding in severely immunocompromised patients with COVID-19

We identified various Omicron sublineages of SARS-CoV-2 in the patients included in this study (Supplementary Figure S2). In both the severely and the moderately immunocompromised groups, BA.5 was the predominant lineage, whereas BA.1 was the most frequently detected lineage in the immunocompetent group. In the severely immunocompromised group, two Delta variant cases were included, with one case (CID-024-1) showing persistent Delta variant infection through May 2022. Minor sublineage shifts were observed in three cases in the severely immunocompromised group and one in the immunocompetent group; in all other cases, viral lineages remained stable throughout the observation period. Phylogenetic analysis of 71 immunocompromised cases (severe and moderate groups) demonstrated distinct clustering patterns for each case (Figure 3a), consistent with independent within-host evolution.

**Figure 3.**
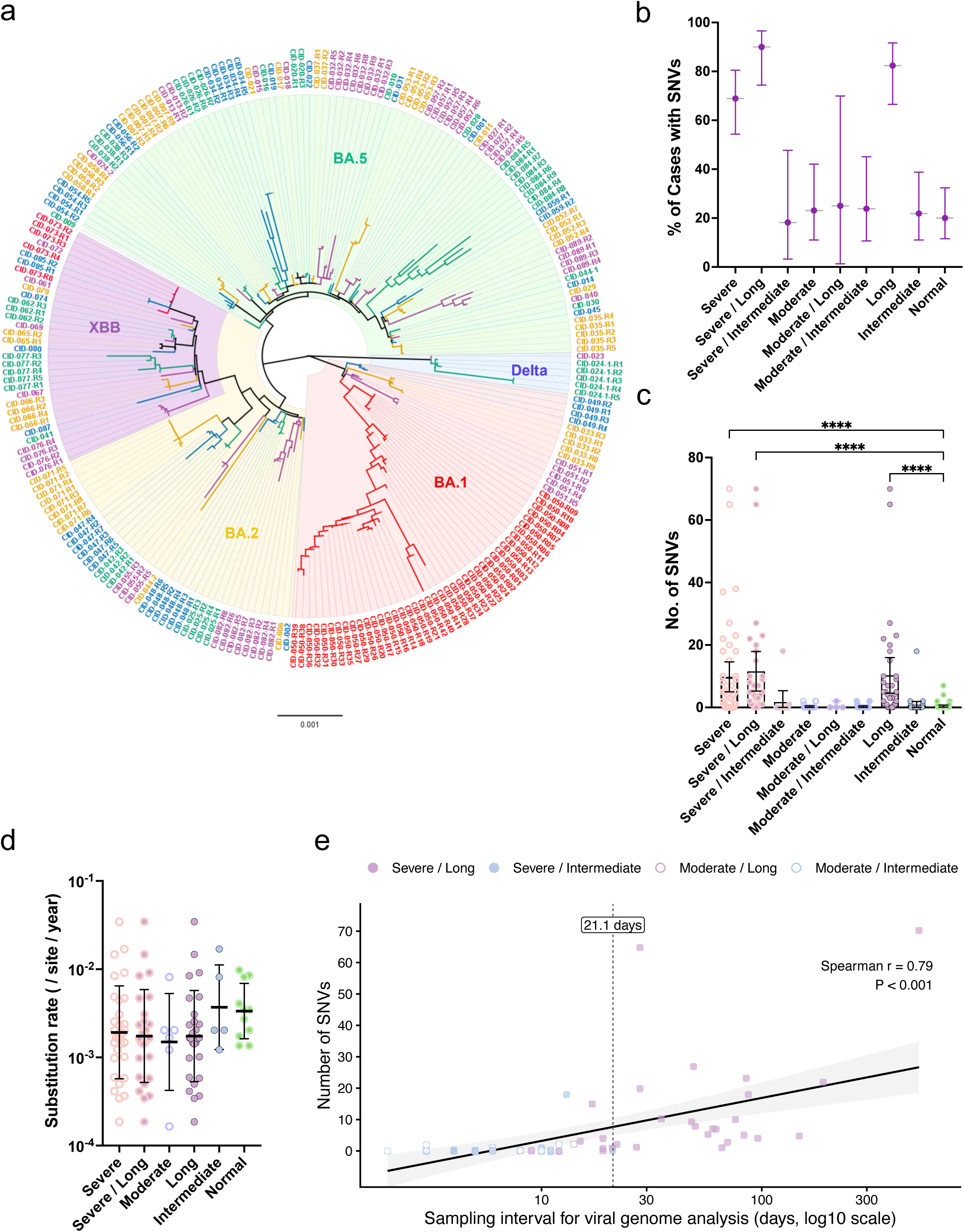
Within-host single-nucleotide variants (SNVs) and substitution rates according to immunocompromised status and model-based shedding stratification. (a) Maximum-likelihood phylogenetic tree of SARS-CoV-2 genomes obtained from 71 immunocompromised cases (Severe and Moderate groups). For cases without detectable intra-host mutations, only one representative sequence is shown (e.g., CID-XXX). For cases with detectable intra-host mutations, sequences from longitudinally collected samples are shown (e.g., CID-XXX-RX). (b) The proportion of cases with detected SNVs (95% confidence intervals) in the immunocompromised status groups (Normal, Moderate, Severe), the model-defined shedding duration groups (Intermediate vs Long), and the combined stratification groups by immunocompromised status and shedding duration. (c) Total SNV number per case across groups stratified by immunocompromised status (Normal, Moderate, and Severe), model-defined shedding duration (Intermediate vs Long), and the combination of immunocompromised status and shedding duration (mean with 95% confidence intervals). Statistical significance is indicated by asterisks (**** P < 0.0001). (d) Substitution rates (substitutions per site per year) in severely immunocompromised, long shedding, and severely immunocompromised/long shedding status groups. Data are presented as dot plots with geometric mean and geometric SD. (e) Relationship between observation period and accumulation of intra-host SNVs. Scatter plot showing the number of detected single-nucleotide variants (SNVs) in each case against the observation period (days from symptom onset). Colored symbols indicate clinical groups (Severe–Long, closed purple circle; Severe–Intermediate, closed light blue circle; Moderate–Long, open purple circle; Moderate–Intermediate, open light blue circle). Black line shows linear regression fits for all cases with 95% confidence intervals.

In the severely immunocompromised group, 31 of 45 cases (68.9%) exhibited single-nucleotide variants (SNVs) during upper respiratory virus shedding, a markedly higher proportion than in the moderately immunocompromised and immunocompetent groups (Figure 3b). The total number of SNVs was also significantly greater in the severely immunocompromised group than in the moderately immunocompromised and immunocompetent groups (Figure 3c). To determine whether this elevated mutation burden reflected immunocompromised status per se or was driven by prolonged virus shedding, cases were further stratified according to the model-defined virus shedding duration. The proportion of cases with detected SNVs was significantly higher in cases with the long shedding phenotype than in those with the intermediate phenotype (Figure 3b), as was the total number of SNVs (Figure 3c). Furthermore, combined stratification based on both immunocompromised status and virus shedding duration demonstrated that both the proportion of cases with detected SNVs and the total number of SNVs were greatest in cases with long shedding phenotype within the severely immunocompromised group, compared with all other combined strata (Figure 3b, c). To assess whether the higher mutation counts in these groups reflected an accelerated rate of mutation introduction rather than simply extended observation time, the substitution rate was calculated as the number of detected SNVs divided by the observation period. However, no significant differences in substitution rates were identified across any of the stratified groups (Figure 3d, Supplementary Figure S3), suggesting that the greater mutation burden observed in the cases with the long shedding phenotype reflects the duration of virus shedding rather than an intrinsically elevated mutation rate. Notably, SNV accumulation remained limited in moderately immunocompromised cases with the long shedding phenotype and in severely immunocompromised cases with the intermediate shedding phenotype (Figure 3c), indicating that neither prolonged shedding nor severe immunocompromise alone was sufficient to drive substantial within-host viral evolution. Additionally, the number of detected SNVs increased with virus shedding duration (Spearman’s ρ = 0.79, P < 0.001; Figure 3e). In a linear model including group-by-time interaction terms, no significant differences in SNV accumulation rates were observed across groups (all interactions P > 0.7).

Taken together, these findings suggested that mutation burden was strongly associated with the long shedding phenotype in addition to severe immunocompromise rather than with immunocompromised status alone.

Importantly, these findings raise the possibility that a practical threshold of infectious virus shedding duration may exist, beyond which within-host viral diversification becomes substantial. In the present cohort, cases with the intermediate shedding phenotype exhibited minimal accumulation of SNVs, even among severely immunocompromised individuals, whereas cases with the long shedding phenotype showed a marked increase in mutation burden. This suggests that maintaining infectious virus shedding duration below a certain threshold may be associated with reduced viral diversification. Although the precise cutoff requires further validation, our data suggest that limiting the duration of infectious virus shedding to approximately 21.1 days (95% CI: 18.9 – 25.6) may substantially reduce the risk of mutation accumulation (Supplementary Figure S4 and S5). From a clinical perspective, this cutoff provides a potentially actionable target for antiviral intervention, as shortening the duration of infectious virus shedding—rather than solely improving clinical symptoms—may be critical for limiting within-host viral evolution.

### Clinical and public health relevance of spike mutations during prolonged virus shedding

Longitudinal measurements of anti–SARS-CoV-2 spike antibody levels demonstrated detectable increases following infection and/or therapeutic antibody administration in severely immunocompromised cases, including those receiving B-cell–depleting therapy (Figure 4a). These findings indicate the presence of humoral immune pressure directed against the spike protein during prolonged virus shedding.

**Figure 4.**
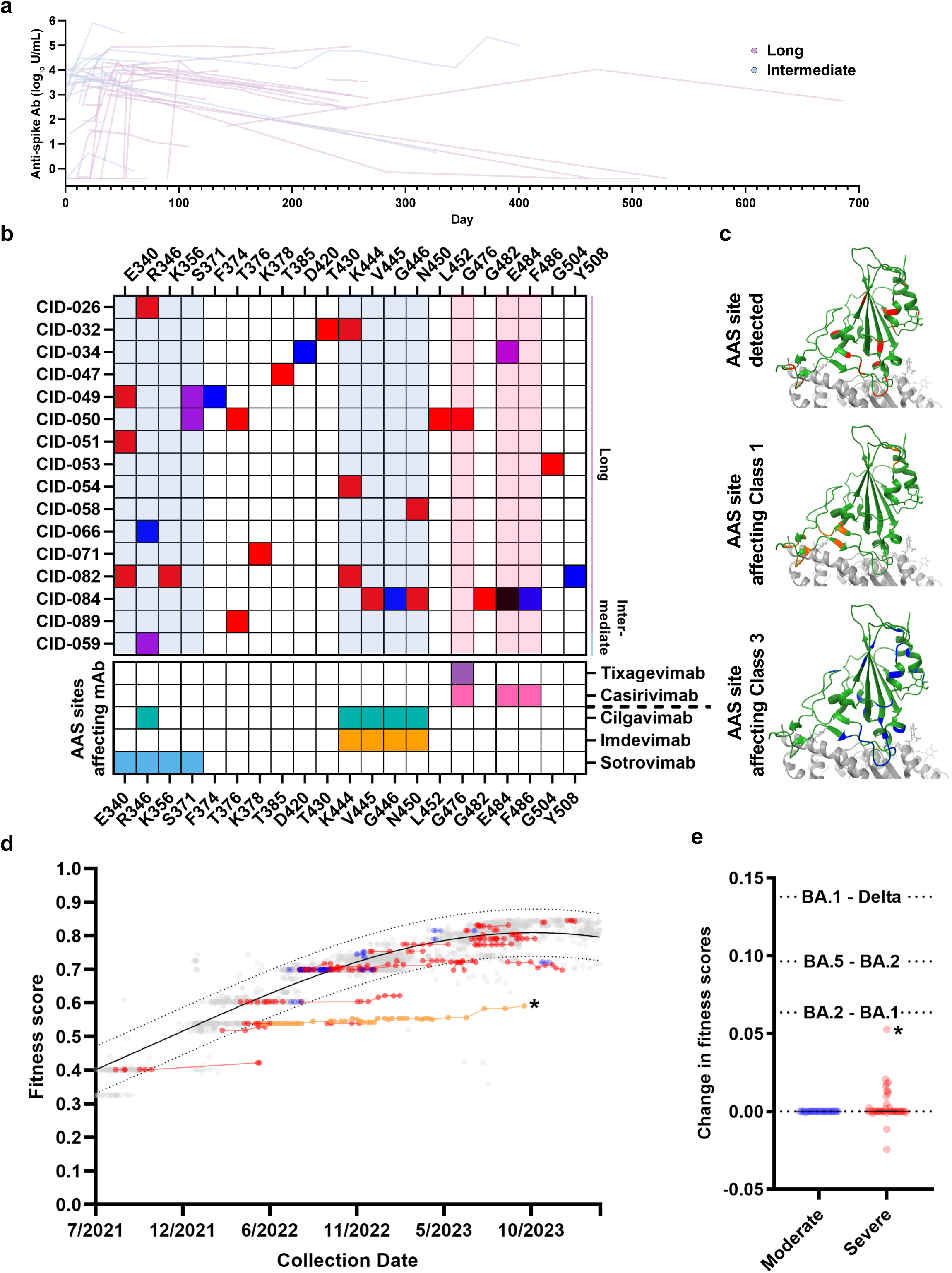
Spike antibody dynamics, resistance-associated mutations, and functional fitness of within-host mutated viruses. (a) Serum anti–SARS-CoV-2 spike antibody levels (log₁₀ U/mL) in Severe cases plotted against days from symptom onset. Each line represents an individual case (purple, Long; light blue, Intermediate). (b) Distribution of amino acid substitutions (AASs) in the receptor-binding domain (RBD) among Severe cases. Mutations are color-coded as follows: red, substitutions from the ancestral strain; blue, reversions to the ancestral residue; purple, changes from one substituted residue to another; black, deletions. The lower panel summarizes the detected AASs and their known resistance associations with therapeutic monoclonal antibodies (mAbs); Class 1 mAbs (above the dashed line), Class 3 mAbs (below). (c) Structural mapping of AAS sites detected in the Severe group (left, red), AASs associated with Class 1 mAb resistance (middle, orange), and those associated with Class 3 mAb resistance (right, blue), displayed on the SARS-CoV-2 RBD (green) in complex with angiotensin-converting enzyme 2 (ACE2; gray). Overlapping positions between observed mutations and Class 3 resistance sites are evident. (d) Fitness scores of SARS-CoV-2 strains circulating in the Japanese community (gray), with a nonlinear regression curve (solid line) and 95% prediction interval (dashed line). Overlaid are the fitness scores of viruses from the Severe group (red/orange) and Moderate group (blue), plotted by sample collection date. The trajectory of CID-050, the case with the longest genome observation period, is highlighted in orange (*). (e) Change in spike protein fitness score from the first to last sampling point in each Severe and Moderate case. For reference, fitness score changes of major community-circulating lineages during the study period are also shown (calculated based on the median fitness scores of each lineage; Supplementary Table S9). CID-050, which showed the largest observed fitness change, is marked with an asterisk (*).

In this context, amino acid substitutions (AASs) within the RBD, a region critical for viral infectivity and neutralizing antibody recognition, were examined in severely immunocompromised cases with multiple detected SNVs (Figure 4b). AASs were observed at multiple RBD sites, including substitutions from the ancestral sequence, reversions to the ancestral state, and substitutions to alternative amino acids. Several AASs identified in severely immunocompromised cases were associated with the binding footprints of Class 1 (tixagevimab and casirivimab) and Class 3 (cilgavimab, imdevimab, and sotrovimab) monoclonal antibodies, which were administered in these cases during the clinical course (Figure 4b and c). Notably, the use of Class 3 monoclonal antibodies was significantly associated with the emergence of resistance-associated AASs among 37 severely immunocompromised cases (excluding eight cases that had been treated with antibody therapeutics before genome observation, Fisher’s exact test, P = 0.012; Supplementary Table S8). No significant association was observed for the use of Class 1 monoclonal antibodies and other antiviral agents (Supplementary Table S9). Notably, in one case (CID-050), the resistance-associated substitution G476S emerged after administration of a Class 1 monoclonal antibody (tixagevimab).

To estimate the inter-host transmission fitness of viruses harboring within-host spike mutations that emerged in immunocompromised cases with COVID-19, we calculated a score reflecting potential community transmission by using the CoVFit pipeline^16^. The fitness score incorporates factors such as the relative effective reproduction number and the ability to evade neutralizing antibody-mediated immunity. Fitness scores were estimated from spike protein sequences of viruses detected in immunocompromised patients with COVID-19 (moderate and severe groups) and compared with those from community-circulating strains in Japan. The circulating lineages in Japan exhibited an increasing fitness trend over time, whereas viruses with within-host spike mutations showed minimal fluctuation in fitness scores (Figure 4d and Supplementary Table S10). None of the viruses detected in immunocompromised patients exceeded the fitness score range of community-circulating viruses in Japan. Even in the case with the longest genome observation period (CID-050), no substantial increase or decrease in fitness score was observed. In addition, although the initial fitness scores of the viruses in immunocompromised patients were comparable to those of community-circulating viruses, the fitness scores of within-host-evolved viruses were subsequently lower than those detected in the community at the same time point (Figure 4d). When changes in fitness scores between early- and late-infection viruses were calculated for immunocompromised cases with COVID-19, no change in fitness score was detected in any case in the moderately immunocompromised group (Figure 4e). In the severely immunocompromised group, individual cases showed either increases or decreases in fitness score; at the group level, however, no net change in fitness score was observed, consistent with the pattern seen in the moderately immunocompromised group (Figure 4e). Furthermore, even in the severely immunocompromised case exhibiting the greatest increase in fitness score (CID-050), the magnitude of change did not exceed the difference in fitness scores observed between the BA.1 and BA.2 lineages (Figure 4e).

Taken together, these results suggest that, compared with the evolutionary changes that occur during typical inter-host transmission in the community, within-host evolution is less likely to generate mutations that enhance inter-host transmissibility.

### Viral genome-wide distribution of within-host mutations during prolonged virus shedding

Because the functional analyses were confined to spike mutations, we next examined the genome-wide distribution of within-host mutations across the virus genome. Specifically, SNV distribution was analyzed in severely immunocompromised cases with multiple detected SNVs (Figure 5a). Detected SNVs were categorized as non-synonymous, synonymous, or transient (i.e., emerging and subsequently disappearing during follow-up).

**Figure 5.**
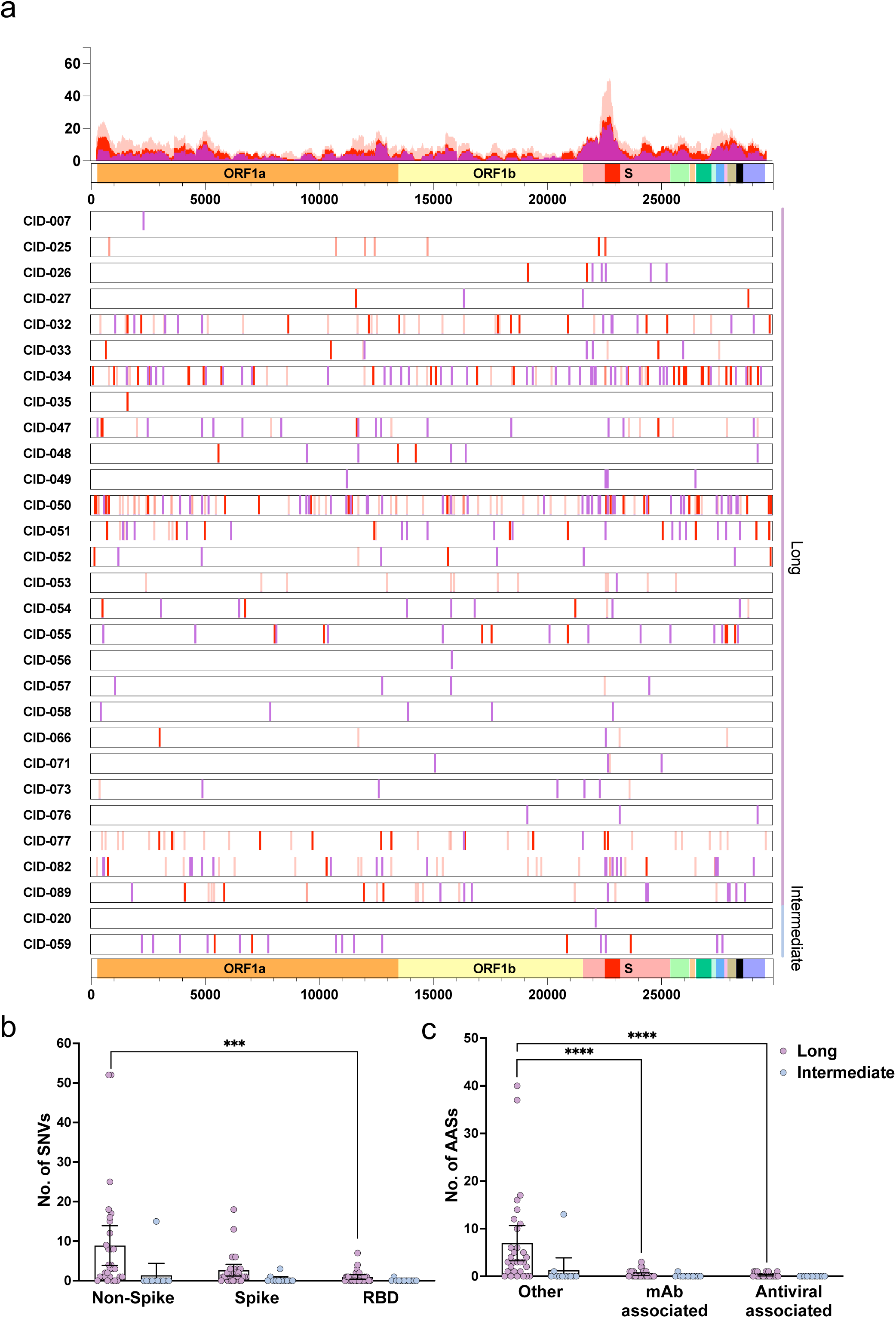
Genome-wide within-host viral diversification in severely immunocompromised cases during prolonged virus shedding. (a) Genomic distribution of SNVs in Severe cases with detectable mutations, categorized as non-synonymous (purple), synonymous (red), and transient mutations (i.e., appeared and disappeared during follow-up; pale red). The spike gene is denoted as “S,” and the receptor-binding domain (RBD) is highlighted in red within the spike region. The top panel presents a sliding window analysis (500-bp window, 10-bp step), showing mutation frequency across the genome: all SNVs (pale red), synonymous + non-synonymous SNVs (red), and non-synonymous SNVs only (purple). (b) Number of SNVs per case in non-spike, spike, and RBD regions in Severe cases stratified by shedding duration. (c) Number of amino acid substitutions (AASs) per case classified according to genomic location: mAb resistance–associated sites, antiviral resistance–associated sites, or other regions. Site classifications refer to genomic positions previously reported to affect therapeutic efficacy, irrespective of whether substitutions conferred resistance in the present study. In (b) and (c), Severe cases are stratified as Long (purple) and Intermediate (light blue). Data are shown as dot plots with overlaid boxplots (median and interquartile range). Asterisks indicate statistical significance (***P < 0.001; ****P < 0.0001).

Across all categories, the spike gene exhibited the highest SNV frequency, particularly within RBD. However, mutations were not confined to the spike gene. SNVs were broadly distributed across the viral genome, including ORF1a, ORF1b, and other structural and non-structural regions. To quantitatively assess this distribution, severely immunocompromised cases were stratified according to virus shedding duration (long versus intermediate), and SNV counts were compared across non-spike, spike, and RBD regions. In cases with the long shedding duration, non-spike SNVs were significantly more numerous than those in the RBD, supporting broader genome-wide diversification in individuals with prolonged virus shedding (Figure 5b). A similar pattern was observed at the amino acid level. When AASs were classified according to their locations as resistance-associated (monoclonal antibody– or antiviral drug–resistance sites) or non–resistance-associated, viruses from cases with the long shedding duration exhibited a significantly greater number of AASs in non– resistance-associated regions than in resistance-associated sites (Figure 5c). In the moderately immunocompromised and immunocompetent groups, SNVs were also detected across multiple genomic regions, including spike (Supplementary Figure S6). However, owing to the limited number of cases and detected mutations, no distinct mutational hotspots were identified in these groups.

The preceding analyses established that genome-wide mutations accumulate randomly in severely immunocompromised cases with the long shedding phenotype over the course of prolonged virus shedding; however, whether this within-host viral evolution follows a directional trajectory remained to be determined. To address this question, we focused on CID-050, the case in this study with the longest virus shedding duration and the greatest number of within-host mutations detected. CID-050 represented the case with the longest genome observation period and the most extensive within-host genomic diversification. The patient had an underlying hematologic malignancy and had received B-cell–depleting therapy (obinutuzumab) within 3 months of COVID-19 onset and was therefore classified as being severely immunocompromised and having the long shedding phenotype. Although the patient’s respiratory function improved after antiviral and corticosteroid treatment, SARS-CoV-2 remained detectable in nasopharyngeal specimens. A total of 44 samples were collected between day 98 and day 686; viral genomes were successfully sequenced from 42 specimens collected between days 98 and 616. Between the first and last sequenced specimens, 70 SNVs and 43 AASs had accumulated, while the viral lineage remained BC.1 (BA.1 sublineage). Bayesian phylogenetic analysis of the 42 viral genomes estimated a median substitution rate of 5.26 × 10^-4^ substitutions/site/year (95% highest posterior density [HPD]: 4.13 × 10^-4^–6.58 × 10^-4^). The unrooted maximum-likelihood phylogenetic tree showed that longitudinally sampled sequences were partitioned into several distinct genetic clusters (Supplementary Figure S7a). In the time-scaled Bayesian phylogenetic tree, the clusters were temporally resolved, revealing three major evolutionary phases over the course of infection, with an additional minor transition in the late phase (Supplementary Figure S7b). Marked temporal shifts were observed between genomes obtained at days 115 and 122, days 186 and 208, and days 504 and 539. Haplotype network analysis further indicated that these transitions were not consistent with simple stepwise accumulation of mutations but instead involved discontinuous genealogical relationships among major haplotype groups (Supplementary Figure S7c). Collectively, these findings indicate that within-host viral evolution in CID-050 proceeded through discontinuous transitions between genetically distinct viral populations rather than along a single phylogenetic trajectory.

These findings demonstrate that during prolonged virus shedding, genome-wide mutations occurred frequently and randomly outside the target sites for neutralizing antibodies and antiviral drugs in severely immunocompromised cases with long shedding durations.

## Discussion

This study provides an integrated clinical, virological, and genomic analysis of SARS-CoV-2 evolution during prolonged upper respiratory virus shedding in immunocompromised patients. By combining longitudinal viral genome sequencing with mathematical modeling of viral RNA loads and infectious virus titers, we demonstrated that the duration of infectious virus shedding in the upper respiratory tract is a key determinant of within-host viral evolution. Patients with prolonged virus shedding accumulated substantially more mutations than did those with shorter shedding durations, even when severely immunocompromised. Although spike mutations were enriched during prolonged virus shedding and were frequently located at binding footprints of therapeutic monoclonal antibodies, mutations were broadly distributed across the viral genome, indicating genome-wide diversification during extended viral replication in the upper respiratory tract.

These findings highlight the importance of limiting the duration of active viral replication in immunocompromised patients and underscore the potential value of aggressive antiviral therapeutic strategies aimed at shortening the duration of upper respiratory virus shedding.

By stratifying patients according to eligibility criteria for monoclonal antibody-based COVID-19 prophylaxis established during the pandemic, we were able to identify severely immunocompromised individuals at elevated risk for extensive viral evolution. This severely immunocompromised group frequently included patients with hematologic malignancies and those recently treated with B-cell–depleting therapies, such as anti-CD20 monoclonal antibodies. The cases in this group exhibited the greatest number of SNVs in their viral genomes.

In one patient with acquired immunodeficiency syndrome (AIDS; CID-084) in this group, 37 SNVs were identified over a 105-day observation period. This observation supports the applicability of the stratification system to other forms of severe immunocompromise beyond hematologic malignancies.

These results are consistent with prior cohort studies highlighting B cell dysfunction and other forms of severe immunosuppression as contributors to persistent virus shedding and within-host diversification^13,14^. Notably, a previous study using the same stratification criteria reported considerably prolonged virus shedding in patients classified as being severely immunocompromised^43^. These findings collectively underscore the clinical utility of this stratification framework in identifying individuals with prolonged virus shedding. Furthermore, the estimated probability of remaining culture-positive was substantially higher in the severely immunocompromised group, highlighting the potential value of risk-stratified approaches to infection control and clinical follow-up in settings where prolonged infectious virus shedding may have practical implications.

However, the classification based on immunocompromised status alone does not fully capture the heterogeneity in viral evolutionary dynamics observed within the severely immunocompromised group. Our findings indicate that the combination of severe immunocompromise and prolonged virus shedding, rather than immunocompromised status alone, is the primary determinant of within-host viral evolution.

SNVs were predominantly detected in the spike gene, particularly within the RBD, where therapeutic monoclonal antibodies exert their effects. This mutation pattern suggests that selective pressure from neutralizing antibody therapy is a key driver of within-host viral evolution in severely immunocompromised individuals. These observations are consistent with previous reports describing the emergence of resistance mutations under antibody pressure in similar populations^5,6^.

However, analysis of spike protein fitness using a protein language model revealed a clear contrast between within-host viral evolution during prolonged virus shedding and the evolutionary dynamics observed in community-circulating strains. While viral fitness in the general population steadily increased over time—driven by the replacement of lineages with more transmissible variants—viruses carrying accumulated within-host mutations in immunocompromised patients exhibited only modest fitness gains and failed to follow the same upward trajectory as circulating Omicron strains. This pattern suggests that within-host evolution during prolonged virus shedding in severely immunocompromised patients may proceed along evolutionary paths less conducive to epidemic spread. This may reflect the fact that circulating Omicron lineages are already highly optimized for transmission, such that additional mutations within the host are more likely to disrupt, rather than enhance, transmissibility. This may explain why extensive within-host diversification does not necessarily translate into increased population-level transmission.

Despite the limited gain in fitness, extensive viral diversification was observed in CID-050. Phylogenetic analysis revealed parallel evolutionary branches, indicative of coexisting subpopulations evolving independently within the host. Notably, the estimated substitution rate in this case was comparable to that of community-circulating strains (5.83 × 10^−4^ substitutions/site/year [95% CI: 5.56 × 10^−4^–6.11× 10^−4^])^3^, suggesting that the observed diversity likely reflects competition among parallel within-host lineages rather than accelerated mutational pressure.

This interpretation is further supported by the absence of significant differences in substitution rates among the severely immunocompromised, moderately immunocompromised, and immunocompetent groups (Figure 3d). This absence indicates that the higher number of SNVs observed in the severely immunocompromised group is attributable to prolonged virus shedding rather than an accelerated mutation rate. These observations are in line with previous autopsy and longitudinal studies reporting compartmentalized viral populations in persistently infected immunocompromised individuals^44-46^.

Collectively, these findings support the notion that within-host virus evolution in immunocompromised patients is shaped predominantly by stochastic processes, with antibody-based therapeutics representing a notable but limited selective pressure, rather than by adaptations enhancing inter-host transmission. Although prolonged virus shedding in immunocompromised patients has been hypothesized as a source of novel VOCs^10-12^, our results suggest that within-host viral mutations in immunocompromised patients are predominantly characterized by stochastic accumulation proportional to virus shedding duration, and that sustained community transmission may represent a comparatively greater source of mutations capable of generating novel VOCs.

Furthermore, most cases in this study occurred during the Omicron-dominant period, when circulating strains had already acquired key spike protein mutations. Notably, some resistance-associated mutations in the RBD reverted to ancestral residues or transitioned to alternative amino acids (as shown in Figure 4b), potentially limiting further fitness gains. This finding aligns with our observation of stable lineages despite ongoing mutation accumulation.

Importantly, our data suggest the existence of a practical threshold of virus shedding duration beyond which viral diversification becomes substantial. Cases with the intermediate shedding phenotype showed minimal SNV accumulation, even in severely immunocompromised individuals, whereas those with the longer shedding duration exhibited a marked increase in mutation burden. The biological basis of this 21-day threshold warrants further consideration. Given that SNV accumulation increases linearly with shedding duration (Spearman’s ρ = 0.79), the disproportionate mutation burden observed beyond 21 days is unlikely to reflect an intrinsically accelerated mutation rate, as substitution rates did not differ significantly across groups (Figure 3d). Rather, it suggests a qualitative difference in the host’s ability to control virus shedding: patients whose shedding duration remained within approximately 21 days—even among those classified as severely immunocompromised—may retain some residual capacity for virus clearance, whereas those exceeding this threshold appear to have lost effective control of virus shedding, leading to the prolonged and extensive within-host diversification observed here. In other words, the 21-day threshold may serve as an immunological surrogate for the transition from "limited but present" immune control to "effectively absent" immune control for virus shedding. Elucidating the immunological determinants that distinguish these two states—whether humoral, cellular, or innate—represents an important avenue for future research, and may ultimately inform the development of host-directed or immunomodulatory strategies to complement existing antiviral therapies. In addition, these findings imply that limiting the duration of infectious virus shedding to below this threshold may suppress within-host viral evolution and provide a quantitative target for antiviral intervention.

Although no VOCs or fitness-enhanced variants emerged in this cohort, the genome-wide and largely random distribution of accumulated mutations indicates that the risk of generating novel variants of public health concern cannot be excluded. The stochastic nature of this mutational process means that prolonged virus shedding creates an expanding opportunity space for the coincidental emergence of mutations with unpredictable functional consequences, even in the absence of directional selective pressure. In this regard, the 21-day shedding threshold identified in our study may serve not only as a virological benchmark but also as a clinically actionable target. These findings underscore the need for intensive antiviral therapeutic strategies specifically aimed at limiting upper respiratory virus shedding to fewer than 21 days in severely immunocompromised COVID-19 patients—a goal that may require extended treatment regimens, combination antiviral approaches, or both.

From a clinical and public health perspective, our findings suggest that focusing exclusively on spike mutations may underestimate the full extent of within-host viral evolution in immunocompromised patients. Instead, prolonged virus shedding should be recognized as a state that facilitates genome-wide viral diversification, potentially generating a broad spectrum of variants with heterogeneous and difficult-to-predict functional properties. Comprehensive genomic surveillance that extends beyond spike-focused sequencing may therefore be warranted in this population, and further studies are needed to define the optimal antiviral strategies for achieving the shedding duration targets identified here. Equally important is the need for immunological investigations to characterize the host factors that distinguish severely immunocompromised patients who achieve viral clearance within 21 days from those who do not. Such studies should examine not only the degree of B-cell depletion or antibody deficiency, but also T-cell–mediated and innate immune responses, which may contribute to residual viral control even in the absence of robust humoral immunity. A more complete understanding of these mechanisms would provide the biological foundation for rational design of combination strategies that pair antiviral agents with immunological interventions, with the shared goal of limiting upper respiratory virus shedding to below the critical threshold identified in this study. This study has several limitations. First, owing to the clinical nature of the specimens, sequencing depth was sometimes limited, and long-read sequencing was not performed. As a result, we were unable to comprehensively characterize the within-host viral quasispecies or detect haplotype-level recombination. In addition, we carried out our modeling of virus dynamics in a subset of individuals (those with repeated RNA and titer measurements) that only partially overlapped with the sequencing cohort; therefore, the linkage between virus shedding duration and genomic evolution should be interpreted as correlative rather than causal. Second, while insertion and deletion events have been highlighted as important features of SARS-CoV-2 evolution in prior studies^47^, our analysis was not optimized to evaluate these changes in detail owing to technical constraints. Third, our cohort did not include solid organ or hematopoietic stem cell transplant recipients, which may limit the generalizability of our findings to certain subpopulations of immunocompromised patients. Finally, our conclusions regarding inter-host transmission fitness were derived from *in silico* fitness scores and should be interpreted accordingly.

In conclusion, our findings identify prolonged virus shedding in severely immunocompromised patients as the central driver of within-host SARS-CoV-2 evolution. While immunocompromised status contributes to this process—particularly in severely immunosuppressed individuals such as those treated with B-cell–depleting therapies—it does not fully explain the observed variability in viral diversification. In this context, prolonged virus shedding emerges as the key condition under which extensive within-host viral evolution occurs. Importantly, the emergence of within-host variants in the setting of prolonged virus shedding does not necessarily translate into increased population-level transmission risk, and infection-control decisions should remain individualized. Furthermore, therapeutic strategies should be designed to shorten the duration of virus shedding, as limiting the period of active viral replication may be critical for curtailing within-host viral evolution.

## Data Availability

The dataset comprises 286 viral genome sequences from immunocompromised patients, registered under BioProject PRJDB20632. The corresponding Run accessions are DRR667741– DRR668026 (see Supplementary Dataset for sample-wise mapping).

## Acknowledgments

We thank all the clinical staff at NCGM, Aichi Cancer Center Hospital, Toranomon Hospital, Kameda Medical Center, Nagoya City University Hospital, Tokyo Metropolitan Toshima Hospital, and Japanese Red Cross Narita Hospital for their dedication to clinical practice and patient care. We also thank Akiko Sataka, Emi Taeda, and Rena Sakamoto at the Department of Pathology, NIID, and Naomi Nojiri, Hazuka Yoshida-Furihata, and Kosuke Murakami at Department of Diagnostic Testing and Technology Research, NIID, for their technical assistance. We further acknowledge the Global Initiative on Sharing All Influenza Data (GISAID) and all data contributors for providing and sharing the SARS-CoV-2 genome data used in this study.

## Author Contributions

Y.H., K.T., No.I., M.I., Na.I., and Tad.S. designed the study. Y.H., K.T., Sh.M., Shu.I., S.S., A.A., T.K., and H.K. performed the experiments. Y.H., K.T., No.I., Y.D.J., J.K., So.M., Shu.I., H.K., N.S., K.H., T.T., M.D.H., and Shi.I analyzed the data. Sh.M., J.K., So.M., Shu.I., H.K., N.S., K.H., and To.S. provided guidance. Y.H., K.T., No.I., and Y. D. J. wrote the original draft of the manuscript. M.I., K.K., Na.I., N.A., Nob.O., M.H., Tak.S., M.M., H.A., N.U., R.H., Y.M., T.A., K.M., and Nor.O. collected clinical samples and contributed to the clinical interpretation of the data. Shi.I and Tad.S. supervised the study and oversaw project administration.

## Supplementary Information

### Supplementary Methods

#### Characterization of prolonged virological shedding dynamics

To support the interpretation of intra-host evolution, we analyzed serial viral RNA load and infectious virus titers using a simple mathematical model to characterize prolonged shedding dynamics. The modeling cohort was defined by the following criteria: (i) symptom onset date was recorded; (ii) viral RNA load measurements were available at two or more time points; (iii) infectious virus titers were measured at one or more time points; (iv) reinfection cases were excluded to avoid complexities related to viral rebound or uncertain timing of reinfection. Using these criteria, we included 70 immunocompromised cases (moderate, 27 cases; severe, 43 cases) and 47 immunocompetent (i.e., normal immune response) cases^1^.

We used an exponential decay model to describe viral clearance over time:

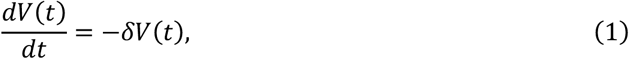

where 𝑉(𝑡) is the viral RNA load (copies/mL) at time 𝑡 since symptom onset and (is the viral clearance rate (1/day). Note that 𝑡 = 0 indicates the date on which symptoms first appeared, and thus 𝑉(0) represents the initial viral RNA load at symptom onset. From Eq.(1), we simply assumed infectious viral shedding dynamics as follows:

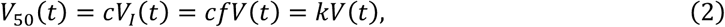

where 𝑉_50_(𝑡) and 𝑉_𝐼_(𝑡) represent the viral titer (TCID50/mL) and the infectious viral RNA load (copies/mL) at time 𝑡 since symptom onset, respectively. The parameter 𝑐 means the conversion from infectious viral RNA load to viral titer, and the infectious viral RNA load is assumed to be a constant fraction of total viral RNA load, denoted by 𝑓. We combined these two parameters as the single constant parameter 𝑘.

Parameters δ 𝑉(0), and 𝑘 were estimated using a nonlinear mixed-effect (NLME) model^2^. This approach explains the heterogeneity in the virological dynamics by including both a fixed effect (the common effect among individuals, i.e., population parameter) and a random effect (the individual-level effect) in each parameter. We used the stochastic approximation expectation maximization algorithm to estimate population parameters and the standard deviation of random effects by computing the maximum-likelihood estimator of the parameters, assuming a Gaussian distribution of residuals (i.e., differences between predictions and measurements)^3^. Individual parameters were obtained as empirical Bayes estimates conditional on the population parameters. The estimation procedures were conducted using MONOLIX 2024R1 (www.lixoft.com).

Furthermore, we inferred the time to culture negativity from the estimated virological trajectories. Infectious virus titers were quantified by a TCID50 assay, and the assay-specific lower limit of detection was set to 24 TCID50/mL (approximately the minimum titer yielding cytopathic effect in at least one well). In our virological trajectories (Eq.(2)), viral titers below the threshold were treated as below the limit of detection (i.e., not culture-positive). For each individual, time to culture negativity was defined as the first time 𝑡^∗^at which 𝑉_50_(𝑡^∗^) dropped below the limit of detection. We then fitted a lognormal distribution to the resulting time-to-event data by maximum-likelihood estimation (MLE). The likelihood for the distribution parameters 𝜇 and 𝜎 given the observations 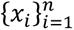 is:

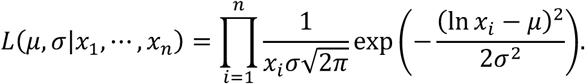

#### Model-based classification of immunocompromised patients by shedding duration

To further characterize heterogeneity in shedding dynamics among immunocompromised cases, we classified these cases using model-estimated features of shedding duration. For each immunocompromised case, we derived two patient-specific quantities from the fitted virological trajectories: the time to RNA negativity and the time to culture negativity. The time to RNA negativity was defined as the first time point at which the predicted viral RNA load fell below the lower limit of detection of the RT-qPCR assay, and the time to culture negativity was defined as the first time point at which the predicted viral titer fell below the assay-specific limit of detection for viral isolation, as described above.

These two features were used as input variables for hierarchical clustering of the 70 immunocompromised cases. Prior to clustering, features were standardized. Euclidean distance was used to quantify pairwise dissimilarity, and hierarchical clustering was performed using Ward’s minimum variance method. Based on the resulting dendrogram, cases were classified into two groups: “intermediate” (i.e., an intermediate duration of virus shedding) and “long” (i.e., a longer duration of virus shedding). These model-based groups were then used for downstream comparisons of viral RNA load at symptom onset, estimated shedding durations, and Kaplan–Meier analyses of RNA positivity and culture positivity.

Furthermore, to explore a practical threshold separating the two model-defined shedding phenotypes, we performed logistic regression analysis using the estimated time to culture negativity as a predictor and the shedding phenotype (intermediate versus long) as the outcome. The optimal cutoff was determined using the Youden index from receiver operating characteristic analysis. The 95% confidence interval of the cutoff was estimated using a bootstrap approach.

#### Retrieval of SARS-CoV-2 sequences from the Japanese community

SARS-CoV-2 genome sequences from Japan were retrieved from the Global Initiative on Sharing All Influenza Data (GISAID). Sequences were filtered using the following criteria: human host; collection date from July 1, 2021, through February 29, 2024; designation as “original”; complete genome sequences; complete collection date information; and availability of patient status metadata. After quality filtering, a total of 4,623 sequences were included in the community reference dataset (Supplementary Dataset).

### Supplementary Results

#### Duration of SARS-CoV-2 viral RNA positivity and infectious virus shedding in the moderately and severely immunocompromised groups

In the moderately immunocompromised group, only a single case (3%) had positive RNA testing results beyond 20 days after illness onset. In contrast, 37 cases (71%) in the severely immunocompromised group had positive results beyond 20 days, including 8 cases (15%) with positive RNA testing results for more than 120 days after illness onset (Supplementary Table S3a).

In the moderately immunocompromised group, one case (3%) tested positive for virus shedding on day 18, and none remained virus-isolation-positive beyond 20 days after illness onset. In contrast, 29 cases (56%) in the severely immunocompromised group continued to shed virus beyond 20 days, including 7 (13%) who remained virus-isolation-positive for more than 120 days (Supplementary Table S3b).

#### Duration of SARS-CoV-2 viral RNA positivity and infectious virus shedding in the model-defined groups with intermediate and long shedding durations

In the group with the intermediate shedding phenotype, 4 cases (11%) had positive RNA testing results beyond 20 days after illness onset. In contrast, 29 cases (83%) in the group with the long shedding phenotype remained RNA-positive beyond 20 days, including 7 (20%) with positive RNA testing results for more than 120 days after illness onset (Supplementary Table S3c).

In the intermediate shedding group, all cases tested negative for virus isolation beyond 20 days after illness onset. In contrast, 26 cases (74%) in the long shedding group continued to shed virus beyond 20 days, including 6 (17%) who remained virus-isolation-positive for more than 120 days (Supplementary Table S3d).

#### Association between antiviral agent exposure and resistance-associated substitutions

The association between antiviral agent exposure and the emergence of resistance-associated amino acid substitutions in nsp5 (main protease) and nsp12 (RNA-dependent RNA polymerase) was examined in severely immunocompromised patients based on previously reported resistance-associated sites (Supplementary Table S9).

Analyses were conducted according to exposure status irrespective of timing or duration of administration, as multiple and overlapping treatment courses were common in this cohort.

No significant association was identified between the use of nirmatrelvir, ensitrelvir, or remdesivir and the emergence of resistance-associated substitutions (Supplementary Table S9).

In remdesivir-exposed patients, seven patients developed substitutions at positions previously implicated in remdesivir resistance in nsp12 (V557L, V792A/I, M794I, and E802K/D), whereas no such substitutions were detected in unexposed patients.

For nirmatrelvir and ensitrelvir, no resistance-associated substitutions were detected in either exposed or unexposed patients. Odds ratios were not estimable in instances with zero events in both groups.

## Supplementary Figures and Tables (Supplementary Figure S1-S7, and Supplementary Tables S1–S10)

**Supplementary Figure S1.**
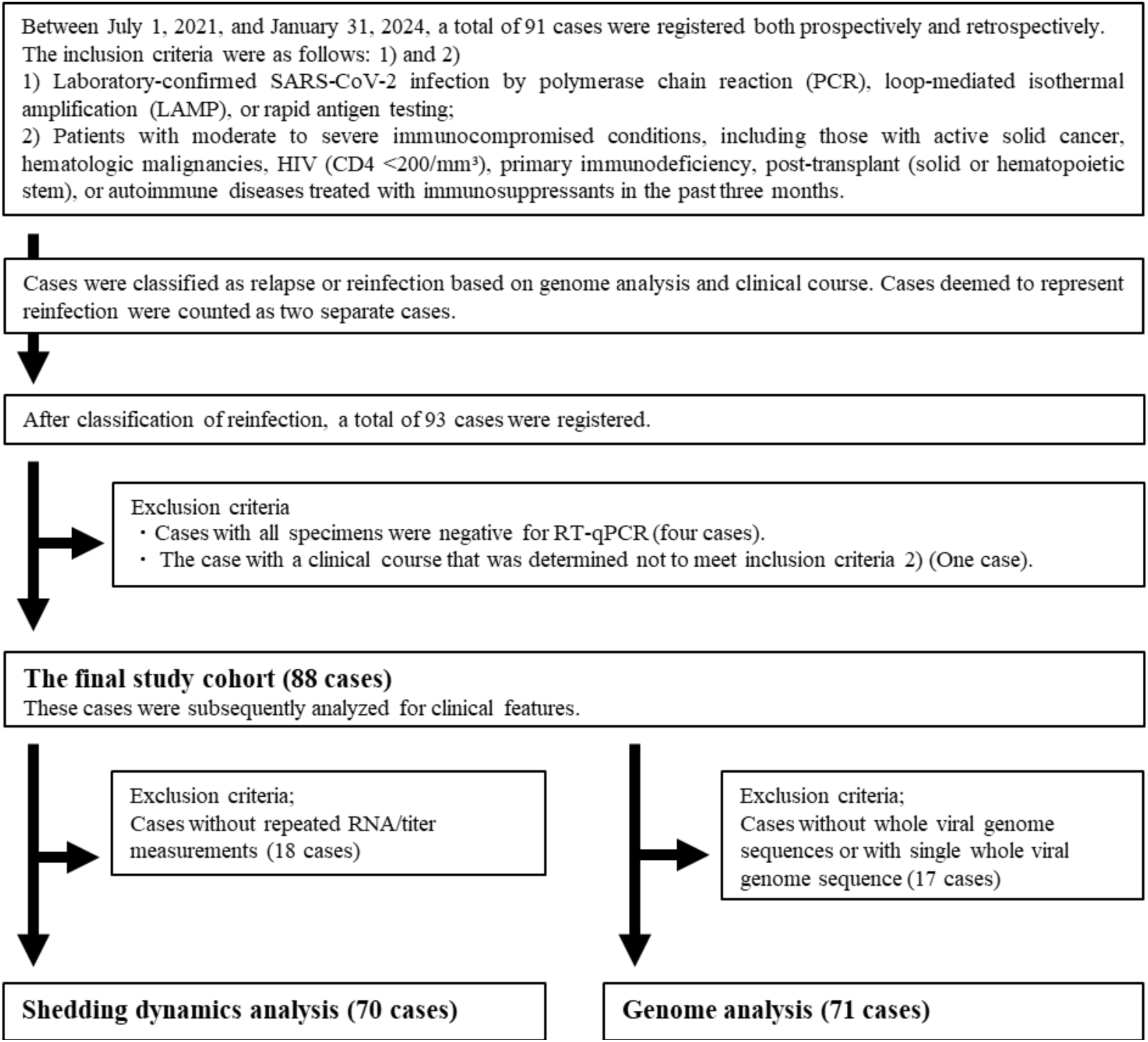
Flowchart of case inclusion.

**Supplementary Figure S2.**
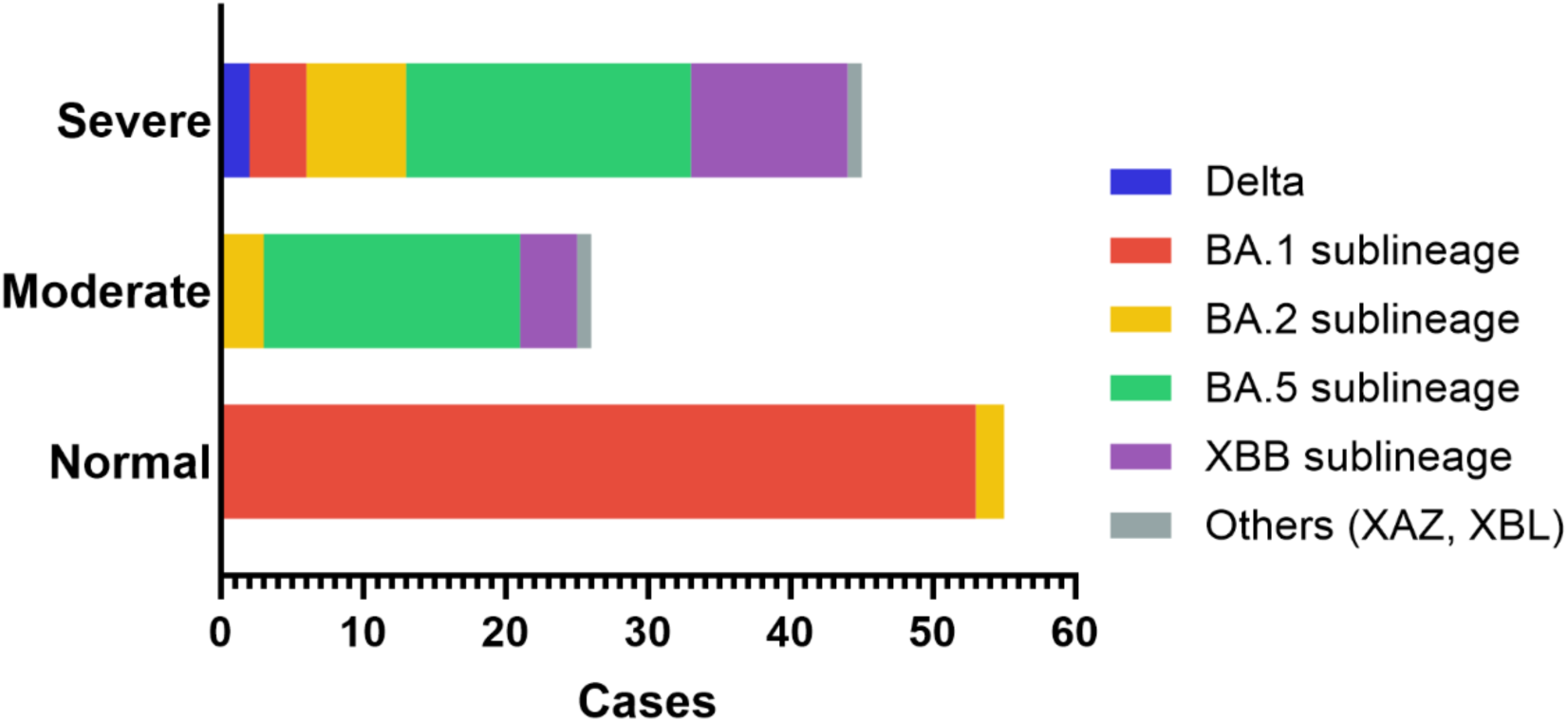
Distribution and proportion of SARS-CoV-2 lineages/clades detected in the severely immunocompromised group (Severe), the moderately immunocompromised group (Moderate), and the immunocompetent group (Normal).

**Supplementary Figure S3.**
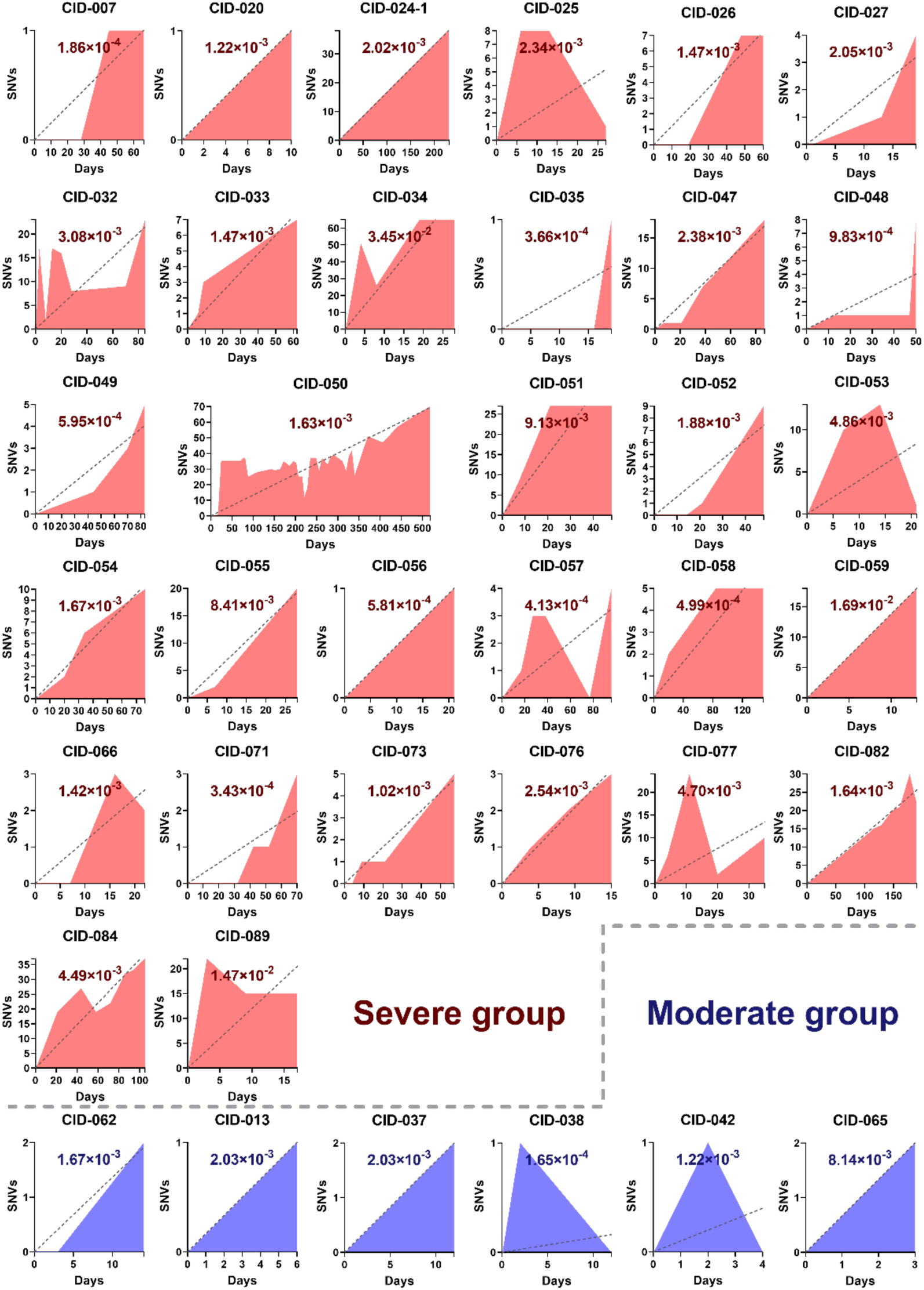
For immunocompromised cases in the Severe and Moderate groups in which SNVs were detected over time, the number of SNVs emerging relative to the first sequenced sample is plotted against the number of days elapsed since that sample. Only cases with detectable SNVs are shown. The slope of a simple linear regression line was used to estimate the substitution rate (substitutions/site/year), assuming a genome length of 29,007 bp based on the SARS-CoV-2 reference genome (MN908947.3). The calculated substitution rate for each case is displayed on the graph. SNVs, single-nucleotide variants; bp, base pairs.

**Supplementary Figure S4.**
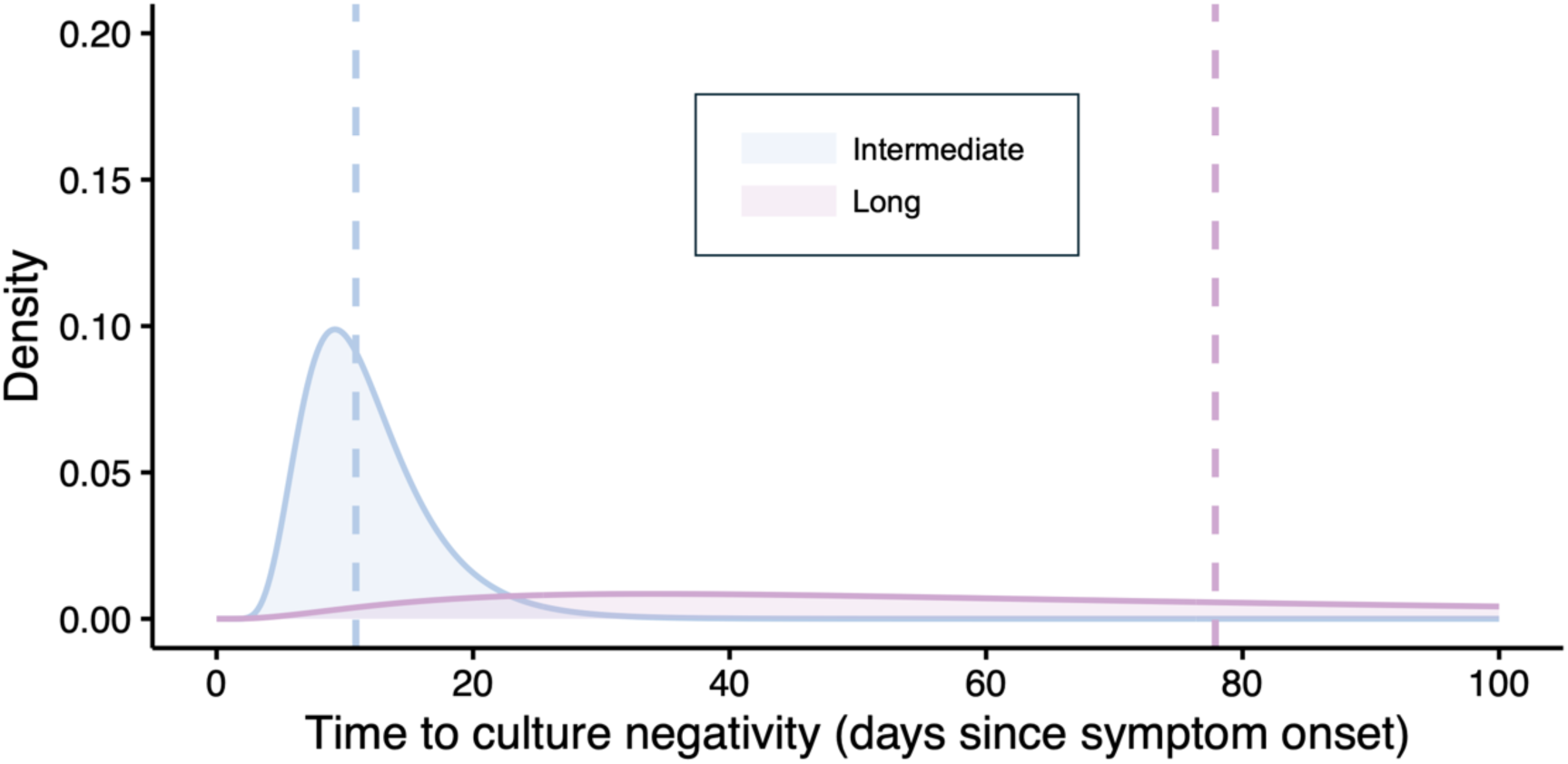
Fitted lognormal probability density functions of the estimated time to culture negativity by model-defined shedding phenotype. Light blue and purple indicate intermediate and long durations of shedding, respectively. Vertical dashed lines indicate the median of each fitted distribution.

**Supplementary Figure S5.**
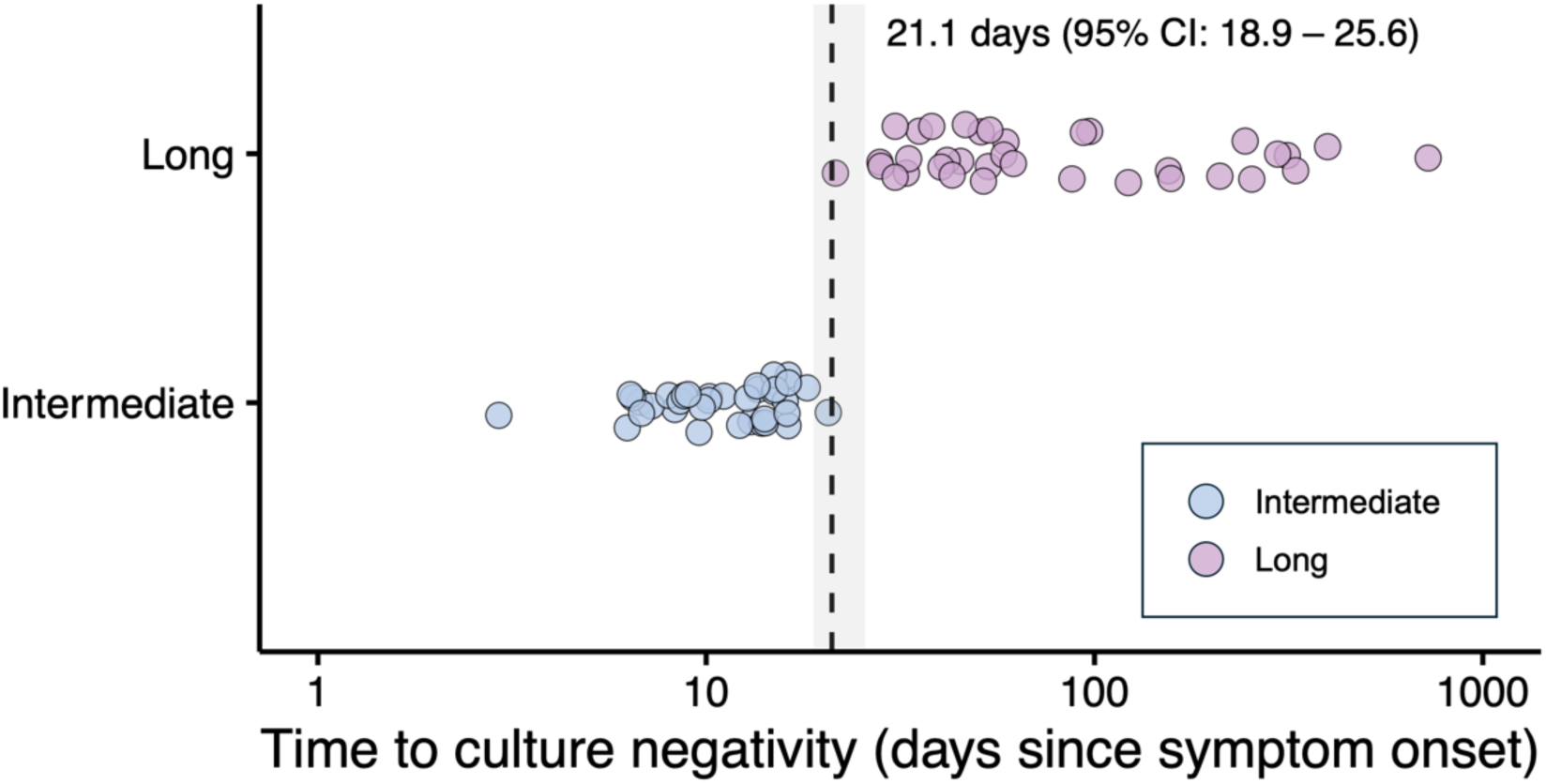
Distribution of estimated time to culture negativity and Youden index-based cutoff for discriminating model-defined shedding phenotypes. Each point represents an individual immunocompromised case, shown on the estimated time to culture negativity (days since symptom onset). Light blue and purple indicate the groups with intermediate and long durations of shedding, respectively. The vertical dashed line indicates the optimal cutoff estimated by logistic regression and receiver operating characteristic analysis using the Youden index. The shaded area indicates the 95% confidence interval of the Youden index via the bootstrap approach.

**Supplementary Figure S6.**
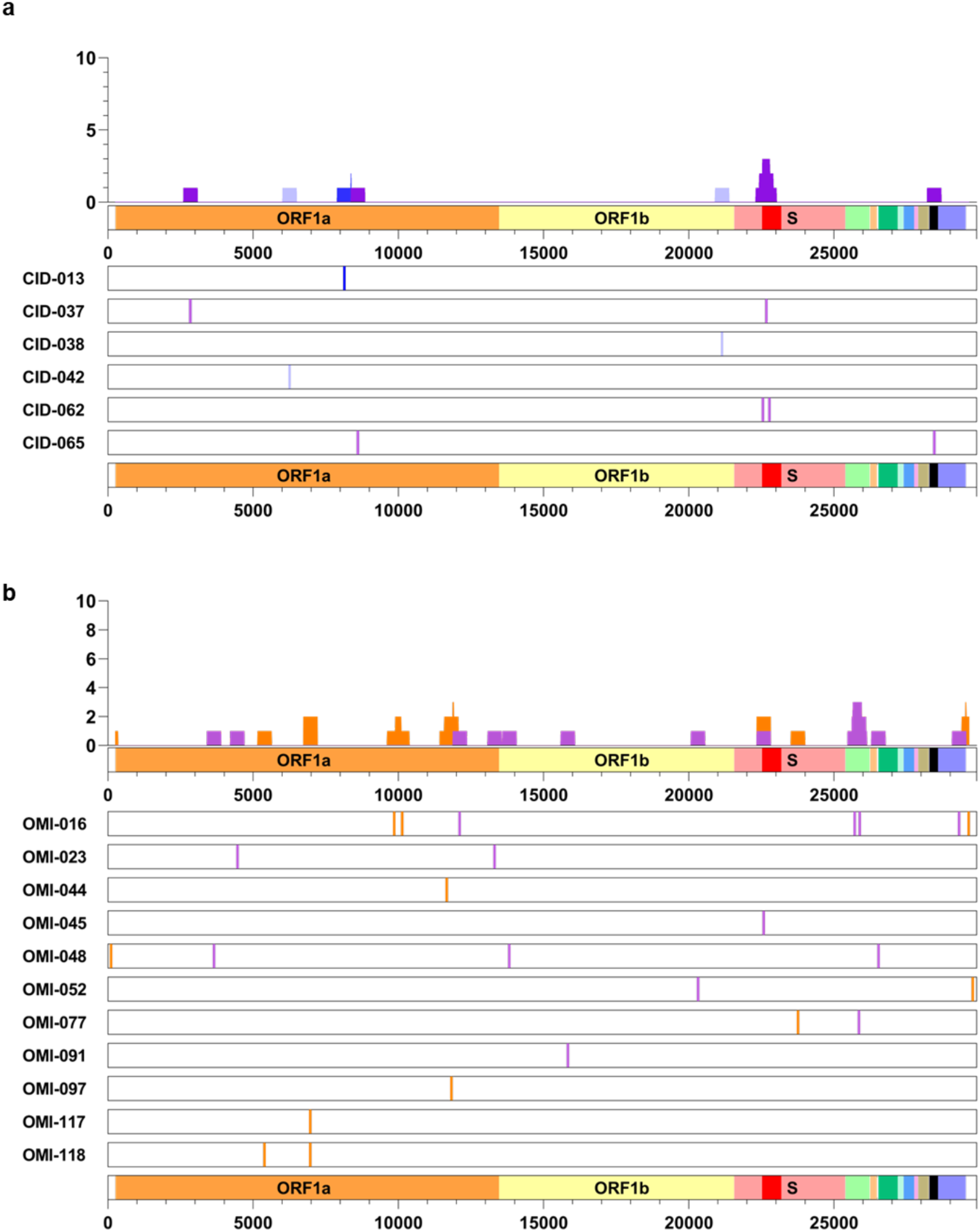
(a, b) Genomic distribution of SNVs in individual cases from the moderately immunocompromised group (a, 6 cases) and the immunocompetent group (b, 11 cases), in which intra-host SNVs were detected. Mutation types are color-coded as follows: non-synonymous SNVs (purple in a and b), synonymous SNVs (blue in a, orange in b), and SNVs that emerged and disappeared during the clinical course (pale blue in a only). The upper panel in each graph presents the results of a sliding window analysis (500-bp window, 10-bp step), showing the total number of each mutation type across cases in which SNVs were detected, plotted along the SARS-CoV-2 genome. SNVs, single-nucleotide variants; bp, base pairs.

**Supplementary Figure S7.**
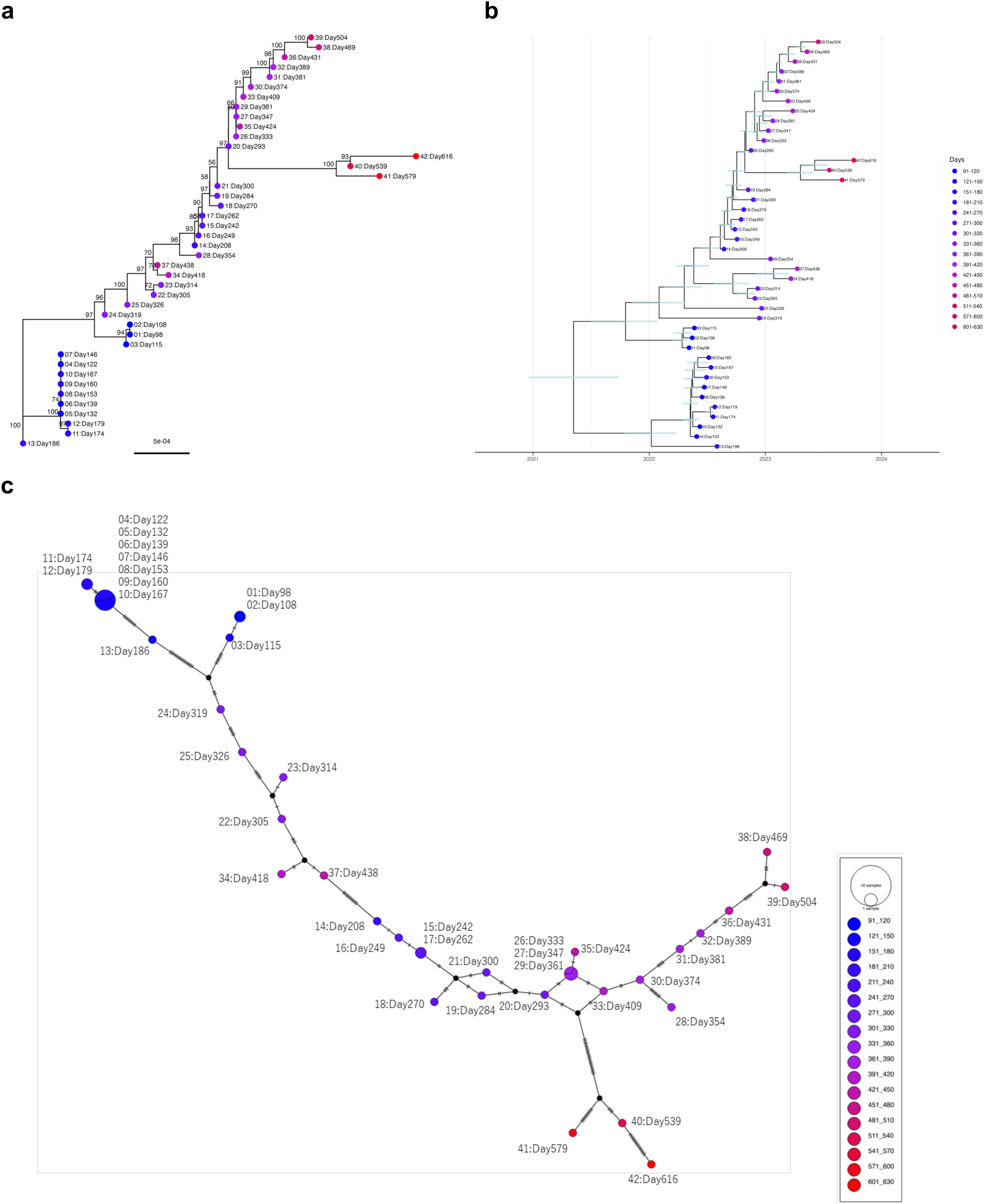
Within-host viral evolution during a prolonged infection (CID-050). (a) Unrooted maximum-likelihood phylogenetic tree of longitudinal viral genome sequences. Bootstrap support values (≥50) are shown at internal nodes. The scale bar represents the number of substitutions per site. (b) Time-scaled phylogenetic tree inferred using Bayesian phylogenetic analysis. The horizontal axis indicates calendar time. Blue bars represent the 95% highest posterior density (HPD) intervals for node heights. (c) Haplotype network constructed using the median-joining method. Each circle represents a unique haplotype, with size proportional to the number of sequences. Hash marks on branches indicate mutational steps. In all panels, tips or nodes represent consensus viral genome sequences labeled with sample number (01–42, in chronological order of sampling) and days from symptom onset. Colors indicate sampling time points, with a gradient from early (blue) to late (red).

**Supplementary Table S1.** Eligibility criteria for tixagevimab/cilgavimab (T/C) in Japan^4^ (Criteria for Inclusion in the Severely Immunocompromised Group, Total n = 53).

| Eligibility criteria | No. of cases (%) |
| --- | --- |
| - Patients with primary immunodeficiency presenting with impaired antibody production or combined immunodeficiency | 8 (15) |
| - Patients within one year of receiving B-cell-depleting therapy (e.g., rituximab) | 32 (60) |
| - Patients receiving Bruton's tyrosine kinase inhibitors | 1 (2) |
| - Chimeric antigen receptor (CAR) T-cell recipients | 2 (4) |
| - Hematopoietic stem cell transplant recipients with chronic graft-versus-host disease or receiving immunosuppressive therapy for other indications | 4 (8) |
| - Patients with hematologic malignancies undergoing active treatment | 18 (34) |
| - Lung transplant recipients | 0 (0) |
| - Patients within one year of receiving solid organ transplantation (excluding lung transplantation) | 0 (0) |
| - Solid organ transplant recipients recently treated with T-cell- or B-cell-depleting agents for acute rejection | 0 (0) |
| - Untreated HIV patients with a CD4 T-cell count of <50 cells/μL | 1 (2) |
| Patients could meet multiple criteria. |  |

**Supplementary Table S2.** Therapeutic agents and resistance-associated amino acid substitution sites investigated in this study.

**a) Monoclonal antibodies (Spike protein; receptor-binding domain)**
| Reported resistance-associated amino acid substitution sites<sup>5-9</sup> |  |
| --- | --- |
| Class 1 |  |
| Tixagevimab | I472, A475, G476, G485, F486, N487, Y489 |
| Casirivimab | C336, C361, K417, Y453, L455, F456, I472, A475, G476, C480, E484, G485, F486, N487, Y489, Q493 |
| Class 3 |  |
| Cilgavimab | C336, R346, C361, E406, Q409, S443, K444, V445, G446, G447, N448, Y449, N450, P463, S494 |
| Imdevimab | C361, N439, N440, S443, K444, V445, G446, G447, N448, N450, P499, P507 |
| Sotrovimab | C336, P337, E340, T345, R346, K356, I358, C361, Y365, Y369, S371 |

**b) Antiviral agents (nsp5 [main protease] and nsp12 [RNA-dependent RNA polymerase])**
| Reported resistance-associated amino acid substitution sites<sup>10-42</sup> |  |
| --- | --- |
| nsp 5 |  |
| Nirmatrelvir | T21, T25, C44, M49, L50, P52, Y54, K90, P108, T135, G138, F140, N142, S144, C160, H163, H164, M165, E166, L167, P168, T169, H172, A173, P184, V186, R188, Q189, A191, Q192, A193, A194, D248, P252, S301, T304, F305 |
| Ensitrelvir | F8, T21, T25, C44, T45, D48, M49, L50 P52, Y54, L57, S144, H16311, H164, M165, E166, L167, P168, P184, Q192 |
| nsp 12 |  |
| Remdesivir | V166, N198, P323, A376, F480, V557, S759, V792, M794, C799, E802 |

**Supplementary Table S3.**
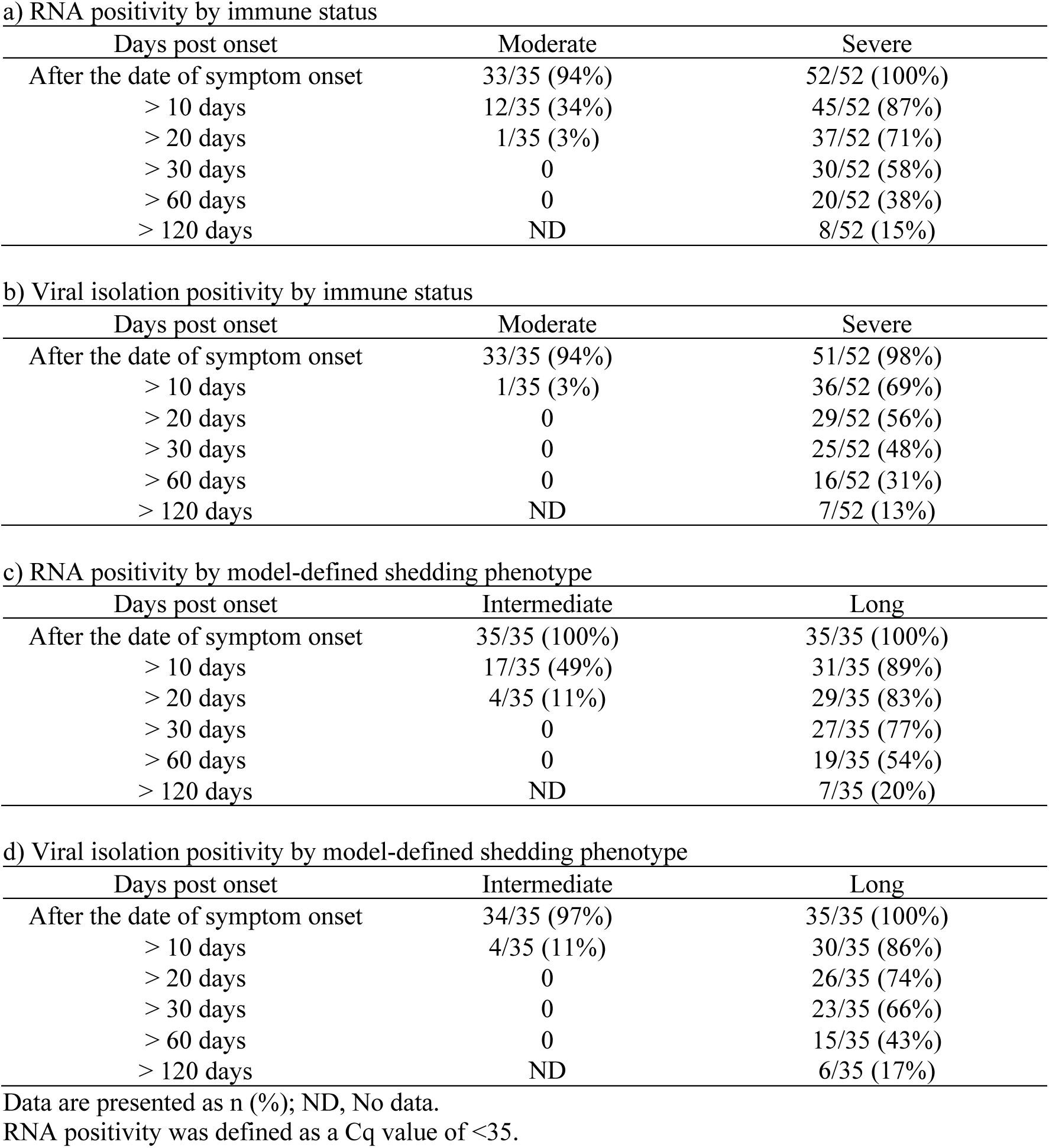
Temporal dynamics of SARS-CoV-2 RNA positivity and viral isolation by immunocompromised status and model-based classification.

**Supplementary Table S4.**
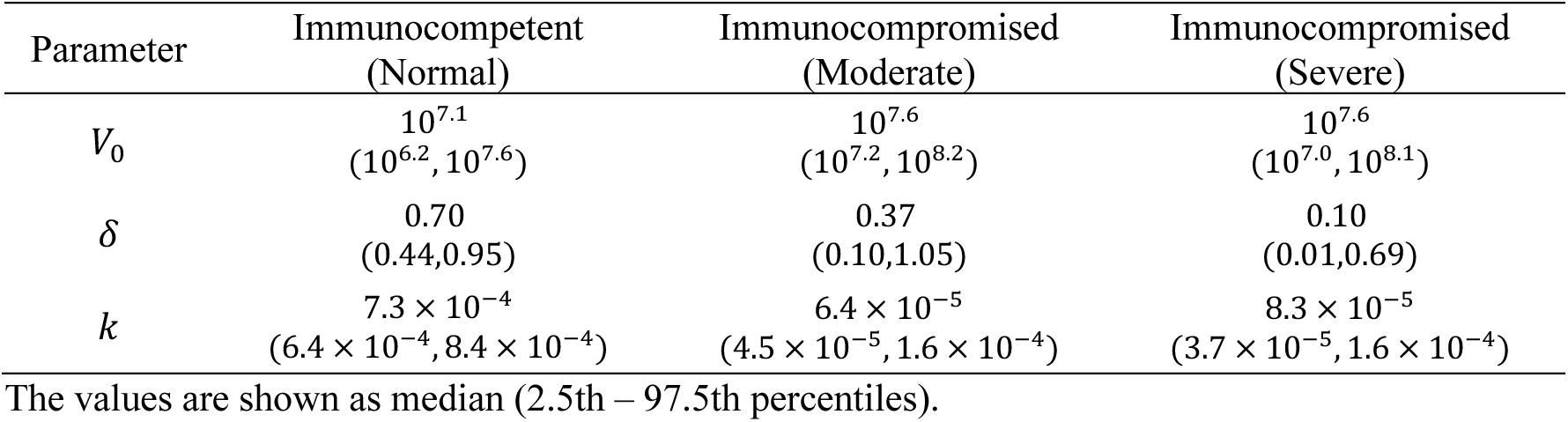
Estimated parameters of virological dynamics model.

**Supplementary Table S5.** Lognormal distribution parameter estimates for time to culture negativity by immunocompromised status.

| Group | Parameter<br>[value (SE)] | Median<br>(days) | Mean<br>(days) | IQR<br>(days) |
| --- | --- | --- | --- | --- |
| Immunocompetent<br>(Normal) | meanlog = 2.15 (0.05),<br>sdlog = 0.31 (0.03) | 8.6 | 9.0 | 7.0 – 10.6 |
| Immunocompromised<br>(Moderate) | meanlog = 2.54 (0.11),<br>sdlog = 0.57 (0.08) | 12.6 | 14.8 | 8.6 – 18.5 |
| Immunocompromised<br>(Severe) | meanlog = 3.89 (0.19),<br>sdlog = 1.21 (0.13) | 49.1 | 102.5 | 21.7 – 111.3 |

SE, standard error; IQR, interquartile range.

**Supplementary Table S6.** Demographic and clinical characteristics of model-defined virus shedding groups in severe immunodeficiency.

|  | Intermediate (n = 12) | Long (n = 31) |
| --- | --- | --- |
| Sex |  |  |
| Men | 9 (75%) [47–91%] | 18 (58%) [41–74%] |
| Age, years | 62 [51–73] | 63 [58–69] |
| Race |  |  |
| Japan | 10 (83%) [55–97%] | 30 (97%) [84–100%] |
| Asia (non-Japan) | 1 (8%) [0–35%] | 1 (3%) [0–16%] |
| Unknown | 1 (8%) [0–35%] | 0 [0–11%] |
| BMI | 22.4 [20.7–24.1] | 20.9 [19.7–22.0] * |
| Comorbidities |  |  |
| Solid cancer | 1 (8%) [0–35%] | 1 (3%) [0–16%] |
| Hematologic malignancy | 11 (92%) [65–100%] | 30 (97%) [84–100%] |
| Autoimmune disease | 0 [0–24%] | 3 (10%) [3–25%] |
| HIV infection | 0 [0–15%] | 1 (3%) [0–16%] |
| AIDS | 0 [0–24%] | 0 [0–11%] |
| Use of immunosuppressants within the past 3 months | 2 (17%) [3–45%] | 9 (32%) [18–51%] # |
| Chemotherapy within the past 3 months (excluding hormonal therapy) | 7 (58%) [32–81%] | 7 (25%) [13–43%] # |
| Evusheld indication |  |  |
| Primary immunodeficiency | 1 (8%) [0–35%] | 5 (16%) [7–33%] |
| B-cell-depleting therapy | 4 (33%) [14–61%] | 25 (81%) [64–91%] |
| Bruton's tyrosine kinase inhibitors | 0 [0–24%] | 1 (3%) [0–16%] |
| CAR T-cell recipients | 0 [0–24%] | 1 (3%) [0–16%] |
| GVHD | 2 (17%) [3–45%] | 2 (6%) [1–21%] |
| Hematologic malignancies undergoing active treatment | 7 (58%) [32–81%] | 8 (26%) [14–43%] |
| Lung transplant | 0 [0–24%] | 0 [0–11%] |
| Solid organ transplantation | 0 [0–24%] | 0 [0–11%] |
| Solid organ transplant recipients with T-cell- or B-cell-depleting agents | 0 [0–24%] | 0 [0–11%] |
| HIV with CD4+ T-cell counts < 50/mm<sup>3</sup> | 0 [0–24%] | 0 [0–11%] |
| Clinical course |  |  |
| Pneumonia on admission | 6 (50%) [25–75%] | 27 (87%) [71–95%] |
| Oxygen therapy | 4 (33%) [14–61%] | 23 (74%) [57–86%] |
| Intensive care | 0 [0–24%] | 9 (29%) [16–47%] |
| Mechanical respiratory support | 0 [0–24%] | 6 (19%) [9–36%] |
| Death | 0 [0–24%] | 10 (32%) [19–50%] |
| Number of COVID-19 vaccinations |  |  |
| 0 | 3 (25%) [9–53%] | 8 (26%) [14–43%] |
| 1 | 0 [0–24%] | 1 (3%) [0–16%] |
| 2 | 2 (17%) [3–45%] | 10 (32%) [19–50%] |
| 3 or more | 7 (58%) [32–81%] | 12 (39%) [24–56%] |
| Treatment |  |  |
| Remdesivir | 11 (92%) [65–100%] | 27 (87%) [71–95%] |
| Molnupiravir | 1 (8%) [0–35%] | 2 (6%) [1–21%] |
| Nirmatrelvir/Ritonavir | 3 (25%) [9–53%] | 15 (48%) [32–65%] |
| Ensitrelvir | 0 [0–24%] | 2 (6%) [1–21%] |
| Tixagevimab/Cilgavimab | 3 (25%) [0–35%] | 17 (54%) [38–71%] |
| Casirivimab/Imdevimab | 0 [0–24%] | 4 (13%) [5–29%] |
| Sotrovimab | 0 [0–24%] | 4 (13%) [5–29%] |
| Tocilizumab | 0 [0–24%] | 8 (26%) [14–43%] |
| Baricitinib | 0 [0–24%] | 6 (19%) [9–36%] |
| Steroids | 4 (33%) [14–61%] | 26 (84%) [67–93%] |
| SARS-CoV-2 lineages |  |  |
| Delta | 0 [0–24%] | 1 (3%) [0–16%] |
| BA.1 sublineage | 0 [0–24%] | 4 (13%) [5–29%] |
| BA.2 sublineage | 0 [0–24%] | 7 (23%) [11–40%] |
| BA.5 sublineage | 5 (42%) [19–68%] | 13 (42%) [26–59%] |
| XBB sublineage | 7 (58%) [32–81%] | 5 (16%) [7–33%] |
| Others | 0 [0–24%] | 1 (3%) [0–16%] |

Data are presented as mean [95% CI, lower limit, upper limit] or n (%) [95% CI, lower limit–upper limit]; BMI, body mass index; COVID-19, coronavirus disease; GVHD, graft-versus-host disease; mAb, monoclonal antibody
\*One case had missing data. #Three cases had missing data.

**Supplementary Table S7.** Lognormal distribution parameter estimates for time to culture negativity by model-defined shedding phenotype.

| Group | Parameter<br>[value (SE)] | Median<br>(days) | Mean<br>(days) | IQR<br>(days) |
| --- | --- | --- | --- | --- |
| Intermediate | meanlog = 2.38 (0.07),<br>sdlog = 0.40 (0.05) | 10.9 | 11.8 | 8.3 – 14.2 |
| Long | meanlog = 4.36 (0.15),<br>sdlog = 0.91 (0.11) | 77.9 | 117.9 | 42.1 – 143.9 |

SE, standard error; IQR, interquartile range.

**Supplementary Table S8.** Association between therapeutic monoclonal antibody use and the emergence of resistance-associated AASs.

| mAb Class | Use | Resistance<br>AAS (+) | Resistance<br>AAS (-) | Odds ratio (95% CI) | *P* value |
| --- | --- | --- | --- | --- | --- |
| Class 1 | Not Used | 0 | 23 | - | - |
|  | Used | 1 | 13 | Infinity (0.1825–Infinity) | 0.3784 |
| Class 3 | Not Used | 1 | 20 | - | - |
|  | Used | 7 | 9 | 15.56 (1.935–181.9) | 0.0118 |

mAb, monoclonal antibody; AAS, amino acid substitution; CI, confidence interval. Resistance-associated AASs were defined as newly acquired substitutions emerging during the genome observation period and were restricted to previously reported resistance sites for each mAb class. Preexisting substitutions and transient variants were excluded from this analysis.

**Supplementary Table S9.**
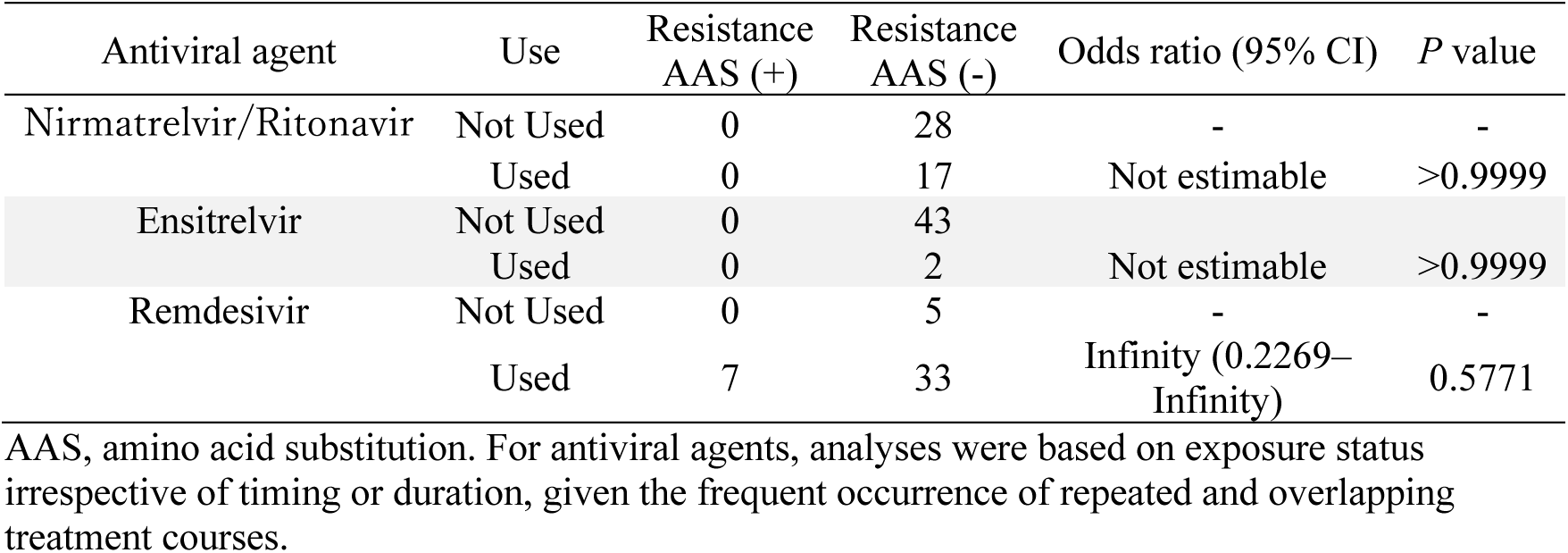
Association between antiviral agent use and the emergence of resistance-associated AASs.

**Supplementary Table S10.** The fitness scores of sequences belonging to the predominant clades circulating during the study period according to sequence data from the Japanese community.

| VOC/Lineage* | Clade_nextstrain* | No. of sequences | Fitness score (median [95% CI]) |
| --- | --- | --- | --- |
| Delta | 21J | 174 | 0.4012 [0.4012, 0.4012] |
| BA.1 | 21K | 362 | 0.5392 [0.5391, 0.5392] |
| BA.2 | 21L | 516 | 0.6028 [0.6028, 0.6029] |
| BA.5 | 22B | 2070 | 0.6993 [0.6993, 0.6993] |

VOC, variant of concern; CI, confidence interval.
\*Classification by Nextclade CLI v.3.8.2<sup>43</sup>.

